# The genetic architecture of changes in adiposity during adulthood

**DOI:** 10.1101/2023.01.09.23284364

**Authors:** Samvida S. Venkatesh, Habib Ganjgahi, Duncan S. Palmer, Kayesha Coley, Laura B. L. Wittemans, Christoffer Nellaker, Chris Holmes, Cecilia M. Lindgren, George Nicholson

## Abstract

Obesity is a heritable disease, characterised by excess adiposity that is measured by body mass index (BMI). While over 1,000 genetic loci are associated with BMI, less is known about the genetic contribution to adiposity trajectories over adulthood. We derive adiposity-change phenotypes from 1.5 million primary-care health records in over 177,000 individuals in UK Biobank to study the genetic architecture of weight-change. Using multiple BMI measurements over time increases power to identify genetic factors affecting baseline BMI. In the largest reported genome-wide study of adiposity-change in adulthood, we identify novel associations with BMI-change at six independent loci, including rs429358 (a missense variant in *APOE*). The SNP-based heritability of BMI-change (1.98%) is 9-fold lower than that of BMI, and higher in women than in men. The modest genetic correlation between BMI-change and BMI (45.2%) indicates that genetic studies of longitudinal trajectories could uncover novel biology driving quantitative trait values in adulthood.

## Introduction

Obesity, the accumulation of excess body fat^1^ which is associated with increased disease burden^2, 3^, has a strong genetic component^4^. The heritability of body mass index (BMI) is estimated to be 40%-70%^4–6^, and genome-wide association studies (GWASs) have implicated over 1,000 independent loci associated with a range of obesity traits^4^. The dynamic process of change in weight over time is also thought to have a genetic component^7, 8^. Recent studies reveal the shifting genetic landscape of infant, childhood, and adolescent BMI, which detect age-specific transient effects by performing age-stratified GWASs^9–11^. Adult twin studies^12–14^ and an electronic health record (EHR)-based population study^15^ indicate that long-term patterns of change in adiposity are heritable and have a distinct genetic component to baseline obesity levels. However, less is known about the specific variants and genes that contribute to patterns of adulthood adiposity change. This paucity of GWASs of long-term trajectories of weight change can be partially attributed to the challenges in building and maintaining large-scale genetics cohorts that follow participants over their lifetime^16^.

Longitudinal data are a key feature of EHRs, whose increased adoption in the clinic and integration into biobanks has powered cost-efficient and scalable genetics research^17, 18^. Despite biases in EHR data, including sparsity, non-random missingness, data inaccuracies, and informed presence, EHR-based genetics studies reliably replicate results from purpose-built cohorts^19–21^. Recent advances in the extraction of phenotypes from longitudinal EHRs at scale show that, as expected^22, 23^, the mean of repeat quantitative measurements can outperform cross-sectional phenotypes for genetic discovery^24, 25^. Repeat measurements further allow for the estimation of longitudinal metrics of trait change, such as trajectory-based clusters^26^, linear slope^27^, and within-individual variability over time^28^, all of which may provide additional information to uncover the genetic underpinnings of disease.

A variety of approaches are available for harnessing the longitudinal component of trajectories in EHR data. Simple models target the gradient of a linear fit over time, such as in a longitudinal linear mixed-effects model framework^28–30^. More complex regression modelling approaches are employed to investigate non-linear changes over time. For example, semiparametric regression models^31^ generate flexible longitudinal patterns from combinations of basis functions, such as B-splines, regularised to induce a suitable degree of temporal smoothness^32–35^. Subgroups of individuals with similar non-linear trajectories are often identified through clustering approaches, with subgroup membership then tested for association with clinical outcomes or genetic variation^36–41^. Although it is possible to fit full joint models that incorporate both genetic data and longitudinal trajectories simultaneously^28^, two-stage approaches wherein summary metrics from models of longitudinal EHRs are taken forward for genetic association analyses are popular for their computational efficiency^27^.

In this study, we leveraged longitudinal primary care EHRs linked to the UK Biobank (UKBB) cohort^42^ to study the genetic architecture of change in adiposity over adulthood. We developed a two-stage analytical pipeline, utilising statistical methods with a history of application in the EHR data context, to derive linear and non-linear trajectories of BMI and weight over time, and to identify clusters of individuals with similar adiposity trajectories. In the second stage, we carried forward the latent phenotypes from these models, which capture both baseline obesity trait levels and change in obesity traits over time, to perform the largest reported genome-wide association analyses for adiposity-change in adulthood. Our results demonstrate the power and added value of EHR-derived longitudinal phenotypes for genetic discovery.

## Results

### Longitudinal data help identify novel genetic signals for obesity

We obtained BMI and weight records for up to 177,098 individuals of white British ancestry with up to 1.48 million measurements in UKBB longitudinal records from general practitioner (GP) and UKBB assessment centre measurements (**Table 1** and **Supp. Fig. 3**). For each individual, we then estimated linear change in BMI or weight over time using a linear mixed-effects model including random intercepts and random longitudinal gradients (**Figure 1A**) within six *strata*—defined as the six pair-wise combinations of the two traits (BMI, weight) with the three sex subsets (women, men, combined sexes).

**Table 1:** Characterisation of obesity trait data in longitudinal records curated from UK Biobank assessment centre visits and linked general practitioner (GP) records. BMI = body mass index, obs. = observation, S.D. = standard deviation, I.Q.R. = inter-quartile range

| Trait | Sex | Number of individuals | Number of obs. | Mean number of repeat obs. (SD) | Mean length of follow-up, years (SD) | Mean age at first obs., years (SD) | Median trait value at first obs. (IQR) |
| --- | --- | --- | --- | --- | --- | --- | --- |
| BMI, kg/m<sup>2</sup> | F | 88,243 (54.4%) | 696,984 | 7.90 (7.34) | 13.7 (6.63) | 48.6 (9.68) | 24.6 (22.2, 27.9) |
| BMI, kg/m<sup>2</sup> | M | 73,965 (45.6%) | 581,161 | 7.86 (7.12) | 12.8 (6.55) | 50.1 (9.59) | 26.1 (24.0, 28.7) |
| Weight, kg | F | 96,625 (54.6%) | 816,885 | 8.45 (8.33) | 13.9 (6.62) | 48.3 (9.63) | 65.0 (59.0, 74.0) |
| Weight, kg | M | 80,473 (45.4%) | 666,258 | 8.28 (7.82) | 12.9 (6.57) | 50.0 (9.57) | 81.6 (73.8, 90.0) |

**Figure 1:**
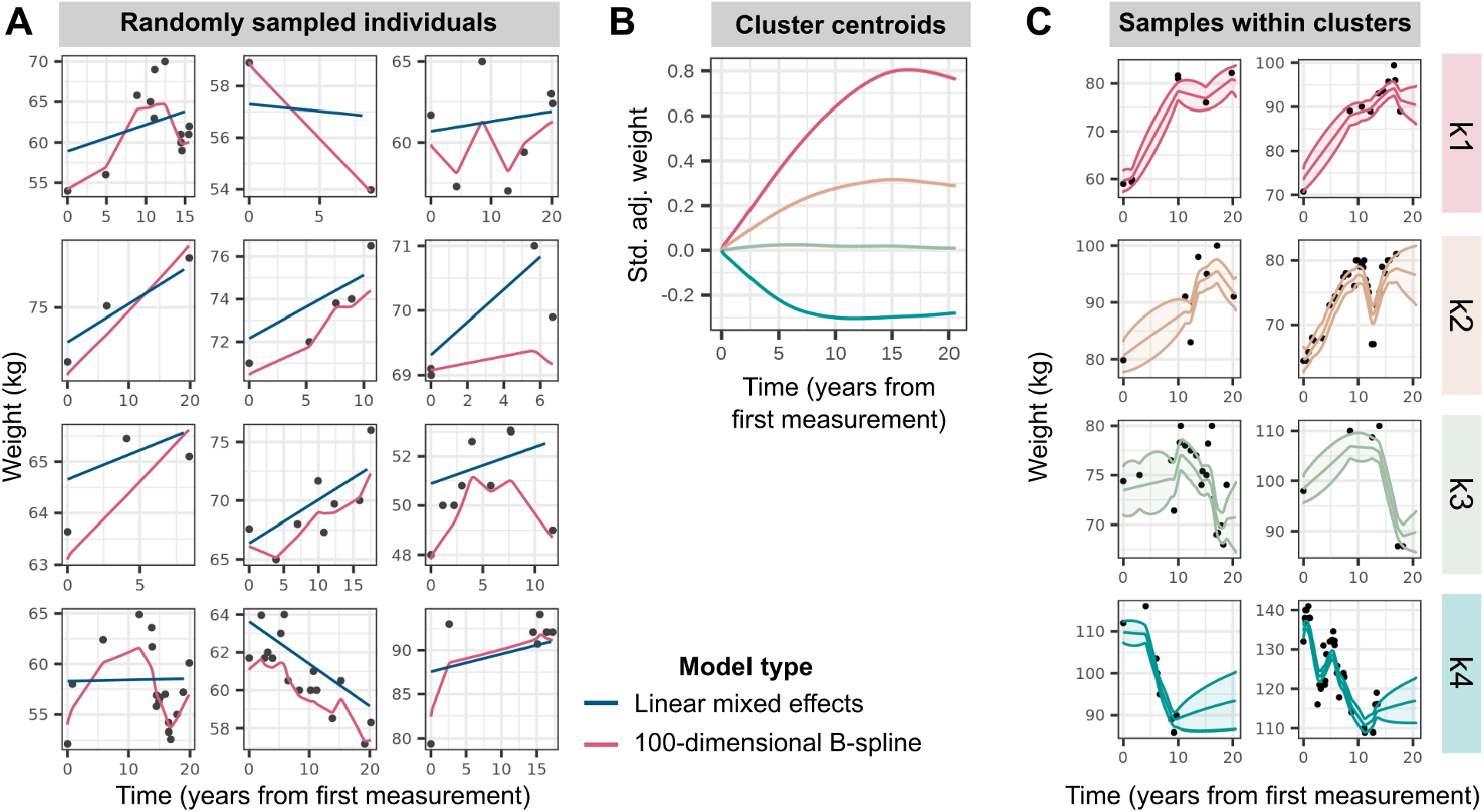
Modelling of longitudinal obesity trait trajectories. **(A)** Weight trajectories over time, measured as years from first measurement, in a random sample of 12 individuals in the sex-combined strata. Black points display observed weight records, with blue and pink lines representing predicted fits from linear mixed effects models and regularised high-dimensional spline models respectively. **(B)** Trajectories of cluster centroids, plotted as standardised (std.) and covariate-adjusted (adj.) weight over time (years from first measurement), for the four clusters determined via partitioning-around-medoids (PAM) clustering with a customised distance matrix (see Methods) constructed from the high-dimensional B-spline coefficients estimated in (A). **(C)** Weight trajectories over time for a random sample of individuals in the 99th percentile probability of belonging to each cluster, as determined by parametric bootstrap. The lines display predicted fits and ribbons represent 95% confidence intervals around the mean fit.

We first investigated whether the individual-level random intercept terms outputted by the longitudinal linear mixed-effects (LME) model, by sharing information across multiple BMI measurements, provided higher statistical power for GWAS than one based on a single, cross-sectional BMI measurement per individual. Despite our GWAS being 4-fold smaller than the largest published analyses^43^, we identify 14 novel loci and refine 53 previously described signals for obesity traits among the 374 unique fine-mapped lead single nucleotide polymorphisms (SNPs) (*P <* 5 *×* 10^−8^) across all strata (**Figure 2A** and (**Supp. Table 2**), see Methods for conditional analysis to classify novel, refined, and reported SNPs^44^). The 53 refined SNPs are conditionally independent of and represent stronger associations (*P <* 0.05) than published SNPs in this population. Together, the refined and novel SNPs explain 0.33% of variance in baseline BMI (in addition to the 2.7% explained by previously published SNPs), and 0.83% of variance in baseline weight (in addition to the 4.7% explained by previously reported SNPs) (**Figure 2B**). We further quantified the power gained from estimating baseline BMI over repeat longitudinal measurements per individual by comparing genome-wide significant (GWS) SNPs from our baseline BMI GWAS to the largest published BMI meta-analysis to date^43^. We observe an increase in median chi-squared statistics of GWS SNPs from either study of between 13.4% (females) to 14.8% (males) in our GWAS over what would be expected from a cross-sectional GWAS of equivalent sample size.

**Figure 2:**
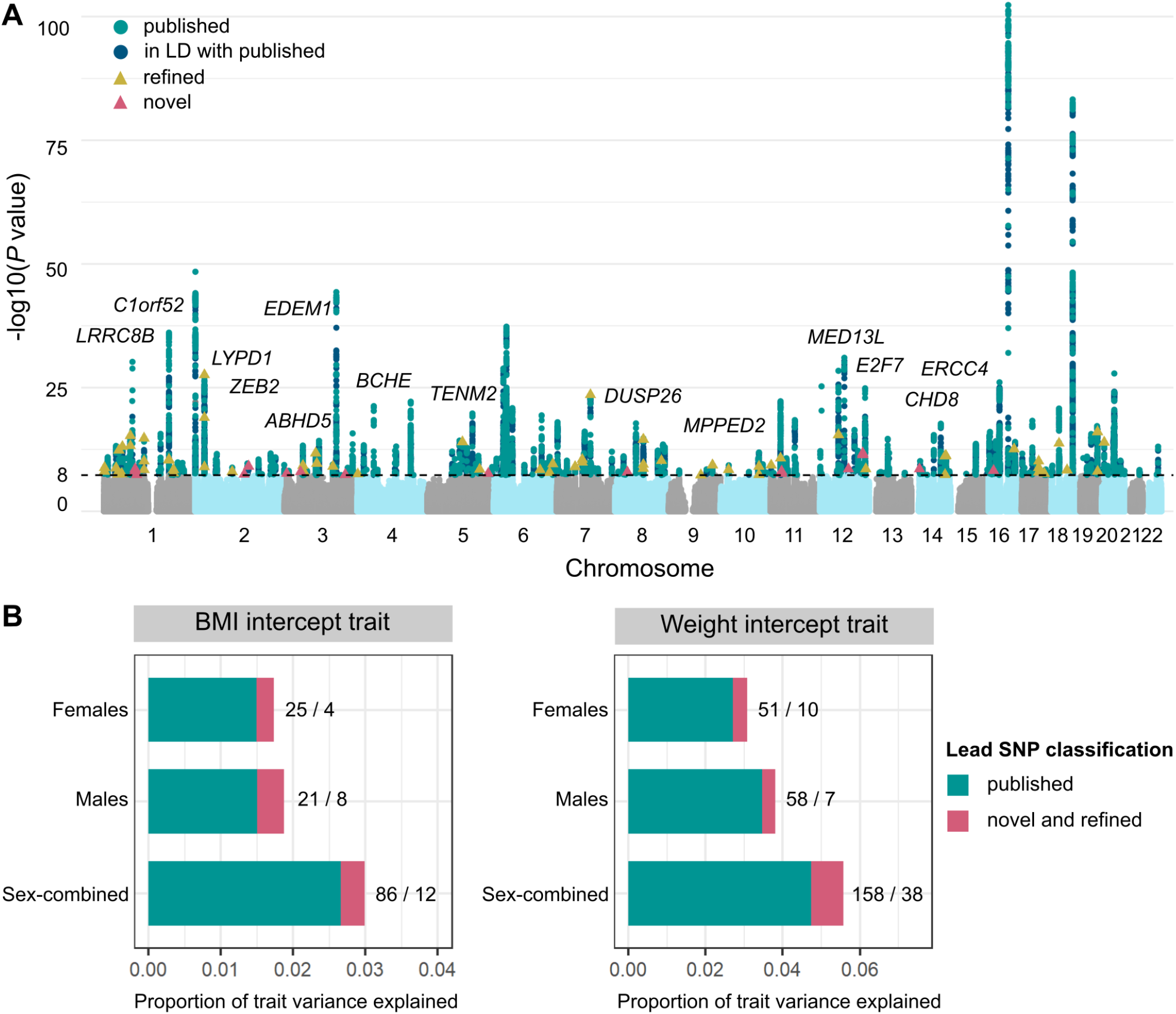
Genome-wide novel and refined SNP associations with baseline obesity estimated over the measurement window for each individual. **(A)** Combined Manhattan plot displaying genome-wide SNP associations with obesity trait (BMI or weight) across female, male, and sex-combined analysis strata. Each point represents a SNP, with GWS SNPs (*P <* 5 *×*10^−8^) coloured in: green for previously published obesity associations, blue for SNPs in linkage disequilibrium (LD) (*r*^2^>0.1) with published associations, yellow for refined SNPs that represent conditionally independent (*P*_*conditional*_ *<* 0.05) and stronger associations with baseline obesity than published SNPs in the region, and pink for novel associations (see Methods^44^). Novel SNPs are annotated to their nearest gene. **(B)** Proportion of variance in baseline BMI and weight that can be explained by the fine-mapped independent lead SNPs in each strata. In green is the proportion of variance explained by previously published obesity-associated variants (and those in LD with these variants), while that explained by novel and refined variants is in pink. The numbers represent the number of lead SNPs in each of these categories (published / refined and novel).

Four of the 14 novel SNPs replicate at *P <* 3.6 *×* 10^−3^ (family-wise error rate (FWER) controlled at 5% across 14 tests using the Bonferroni method) in UKBB assessment centre measurements of cross-sectional obesity in up to 230,861 individuals not included in the discovery GWAS (**Supp. Table 3**). One such novel variant is rs7962636 (*β* = 0.023 standard deviation (SD) increase in expected baseline weight, *P* = 3.7 *×* 10^−12^), whose nearest gene *MED13L* is a transcriptional regulator of white adipocyte differentiation^45^. Twelve of the 14 novel SNPs have a significant (*P <* 3.6 *×* 10^−3^) association with an obesity-related trait in published GWASs that include non-UKBB participants (**Supp. Table 3**). These include rs2861761, whose nearest gene *TENM2* is enriched in white adipocytes^46^, rs13059102, whose nearest gene *BCHE* encodes the liver enzyme butyrylcholinesterase that is associated with obesity via its effect on the ghrelin pathway^47–49^, rs11156978 whose nearest gene *CHD8* is associated with impaired glucose tolerance in mouse knockouts^50^, and rs17794645, whose nearest gene is *ERCC4*, deficiency of which is associated with decreased body weight in mice^51^.

Ascertainment bias in our discovery cohort could arise from the over-representation of heavier participants in EHR data (**Supp. Table 4**)^52^. On average, women with ten or more weight measurements are 8.3 kg (3.7 units of BMI) heavier than their counterparts with 1-3 measurements; for men, this is an 8.2 kg (3.1 units of BMI) difference. However, the BMI intercept metric from our longitudinal data is genetically perfectly correlated with the un-ascertained cross-sectional BMI in Genetic Investigation of ANthropometric Traits (GIANT) 2019^43^ (*r*_*G*_ = 1 and *P <* 1 *×* 10^−16^ in all strata), and 96% of the GWS associations (*P <* 5 *×* 10^−8^) identified in our GWAS have either been reported, or are correlated with reported obesity-associated SNPs in the GWAS Catalog^53^ (**Supp. Table 1**).

### *APOE* missense variant rs429358 is associated with weight loss over time, independent of baseline obesity

To identify genetic variants that affect change in adiposity over time, we performed GWASs for patterns of BMI and weight change adjusted for baseline measurements, defined in two ways. First, we created a linear phenotype from subject-specific random gradients, estimated within a linear mixed-effects model framework, adjusted for corresponding random intercepts, and transformed with a rank-based inverse normal transformation (see Methods). To capture non-linear patterns of temporal change, we additionally modelled longitudinal variation in obesity traits using a regularised high-dimensional B-spline basis^31^ (**Figure 1**). Within each of the six strata, we identified four clusters of individuals using *k*-medoids clustering^54, 55^, representing high gain (k1), moderate gain (k2), stable (k3), and loss (k4) trajectories (**Figure 1** and **Supp. Fig. 5**). We then estimated each individual’s probability of belonging to a cluster based on their posterior non-linear obesity trait trajectory. We performed GWASs on the logit-transformed posterior probabilities of k1, k1 and k2 (high and moderate gain clusters), and k1, k2, and k3 (all but the loss cluster) membership, adjusted for the baseline obesity trait (see Methods).

A common missense variant in *APOE* (rs429358) is associated with decrease in both BMI and weight over time, and lower posterior probabilities of gain-cluster membership in all analysis strata (**Table 2**). Each copy of the minor C allele of rs429358 (minor allele frequency (MAF)=0.16) is associated with 0.060 SD decrease (95% confidence interval (CI)=0.050-0.069, *P* = 8.6 *×* 10^−35^) in expected BMI slope over time and 0.063 SD decrease (0.054-0.072, *P* = 6.0 *×* 10^−42^) in expected weight slope over time (**Figure 3A**). Independent of baseline obesity, carriers of the minor C allele of rs429358 are at lower odds of membership in the high-gain BMI and weight clusters (odds ratio (OR)=0.976, 95% CI=0.97-0.98, *P <* 4.9 *×* 10^−19^), lowering the membership posterior probability from 40% to 39% on average (**Figure 3B**). Although the minor allele of rs429358 is also associated with lower baseline BMI (*β* = 0.015 SD lower BMI intercept, 95% CI=0.0054-0.024) and weight (*β* = 0.011 SD lower weight intercept, 95% CI=0.0029-0.020), these associations do not reach genome-wide significance in any analysis strata (*P >* 0.002).

**Table 2:**
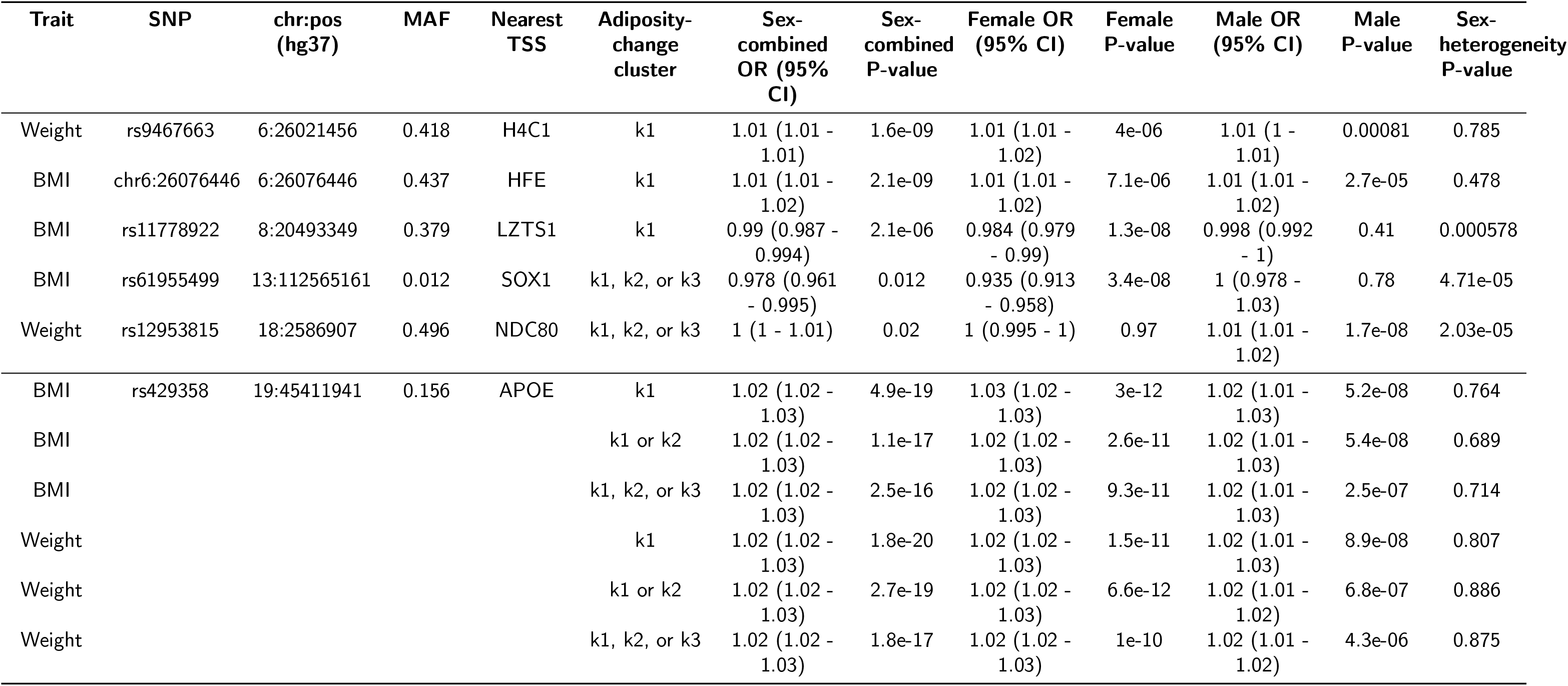
Lead SNPs identified from genome-wide association studies (GWAS) of posterior probability of membership in an adiposity-change cluster (high gain k1, high/moderate gain k1/k2, or high/moderate gain and steady k1/k2/k3), independent of baseline obesity. MAF = minor allele frequency (European-ancestry), TSS = transcription start site, SE = standard error, OR = odds ratio, CI = confidence interval

**Figure 3:**
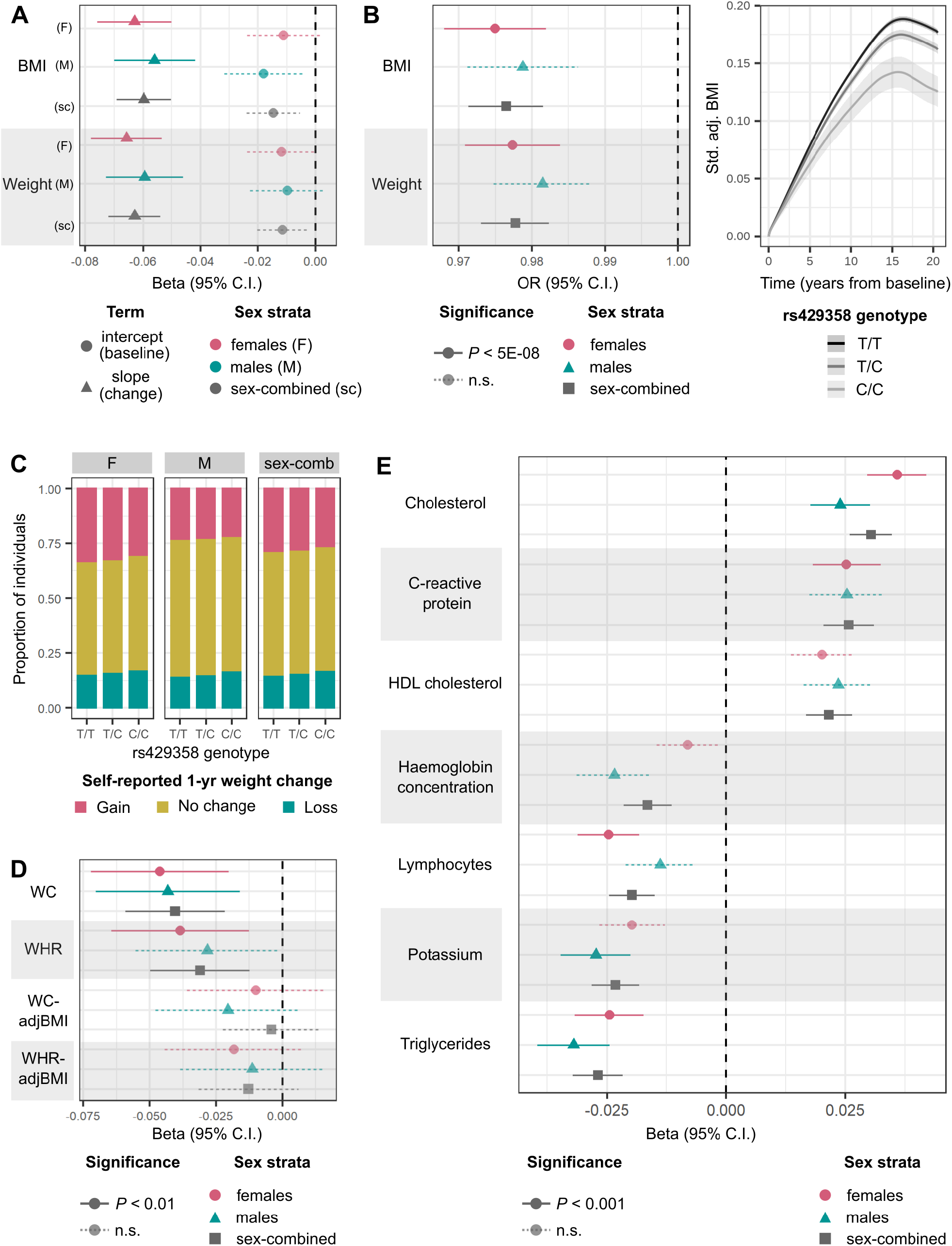
Association of minor C allele of rs429358, missense variant in *APOE*, with various longitudinal phenotypes. **(A)** Effect size (beta) and 95% CI for associations of rs429358 with BMI and weight intercepts or linear slope change over time estimated from linear mixed-effects models in all analysis strata. **(B)** Left: OR and 95% CI for association of rs429358 with posterior probability of membership in the BMI and weight high-gain clusters (k1). Right: Modelled trajectories of standardised (std.) covariate-adjusted (adj.) BMI in carriers of the different rs429358 genotypes. **(C)** Proportion of individuals who self-report weight gain, weight loss, or no change in weight over the past year for carriers of each rs429358 genotype. **(D)** Effect size and 95% CI for associations of rs429358 with slopes over time of waist circumference (WC) and waist-to-hip ratio (WHR), adjusted for BMI (-adjBMI), estimated from linear mixed-effects models. **(E)** Effect size and 95% CI for associations of rs429358 with linear slope change in quantitative biomarkers over time, estimated from linear mixed-effects models. Across all panels, estimates of trait change are adjusted for baseline trait values, and P-values for significance are controlled at 5% across number of tests performed via the Bonferroni method. n.s.=non-significant

The association of rs429358 with adiposity-change phenotypes, across all strata, was replicated at *P <* 5*×*10^−3^ in up to 17,035 individuals in UKBB with multiple measurements of weight and BMI at repeat assessment centre visits who were excluded from the discovery set (**Supp. Table 5**). Based on 301,943 UKBB participants who reported weight change in the last year as “gain”, “about the same”, or “loss”, and who were not included in the discovery GWAS, we found that carriers of each additional copy of the minor C allele of rs429358 are at 0.956 (95% CI=0.94-0.97) lower odds of being in a higher ordinal weight-gain category, independent of their BMI (**Figure 3C** and **Supp. Table 6**). We observe consistent effect direction of the rs429358 association with both estimated and self-reported weight loss over time in individuals who self-identify as Asian (maximum N=8,324 individuals), Black (6,796), mixed (2,681), white not in the white-British ancestry subset (47,174), and other (3,994) ethnicities (see Methods for ancestral group definitions, **Supp. Fig. 1** and **Supp. Table 7**).

Finally, we tested for the effect of rs429358 on change in abdominal adiposity in up to 44,154 individuals of white British ancestry in UKBB who were not in the discovery set, with repeated assessment centre measurements of waist circumference (WC) and waist-to-hip ratio (WHR). Each copy of the C allele is associated with 0.040 SD decrease (95% CI=0.021-0.049, *P* = 2.3 *×* 10^−5^) in expected WC slope over time and 0.031 SD decrease (0.012-0.050, *P* = 1.1 *×* 10^−3^) in expected WHR slope over time, independent of baseline values (**Figure 3D** and **Supp. Table 6**). While the effect direction remains consistent, these associations are no longer significant upon adjustment for BMI (all *P >* 0.1), suggesting that the observed loss in abdominal adiposity over time may represent a reduction in overall adiposity.

The *APOE* locus is a highly pleiotropic region that is associated with lipid levels^56, 57^, Alzheimer’s disease^58, 59^, and lifespan^60, 61^, among other traits^62^. Excluding the 242 individuals with diagnoses of dementia or Alzheimer’s disease in our replication datasets did not alter associations of rs429358 with any of the longitudinal obesity traits (**Supp. Fig. 2**), indicating that they are unlikely to be driven solely by weight loss that accompanies dementia. We additionally performed a longitudinal phenome-wide scan to test for the association of rs429358 with changes in 45 quantitative biomarkers obtained from the UKBB-linked primary care records. Each copy of the C allele is associated with an increase in expected slope change over time of total cholesterol (*β* = 0.030 SD increase, *P* = 6.4*×*10^−12^), C-reactive protein (CRP) (*β* = 0.026, *P* = 9.6*×*10^−7^), and high-density lipoprotein (HDL) cholesterol (*β* = 0.022, *P* = 1.0 *×* 10^−5^), but a decrease in expected slope change over time of triglycerides (*β* = −0.027, *P* = 2.7 *×* 10^−7^), potassium (*β* = −0.023, *P* = 3.9 *×* 10^−6^), lymphocytes (*β* = −0.020, *P* = 4.0 *×* 10^−5^), and haemoglobin concentration (*β* = −0.016, *P* = 1.0 *×* 10^−3^) (FWER controlled at 5% across 45 tests via the Bonferroni method) (**Figure 3E** and **Supp. Table 8**).

### Genome-wide architecture of change in adiposity over time is sex-specific and distinct from baseline adiposity

We identify six independent genetic loci associated with distinct longitudinal trajectories of obesity traits by performing GWAS on individuals’ posterior probabilities of membership in the high gain cluster (k1), high and moderate gain clusters (k1 and k2), or no loss (clusters k1, k2, and k3), adjusted for baseline obesity trait (**Table 2**). This included the *APOE* locus and five signals in intergenic regions. rs9467663 (OR=1.011 for membership in the high-gain weight cluster, *P* = 1.6*×*10^−9^) and chr6:26076446 (OR=1.012 for membership in the high-gain BMI cluster, *P* = 2.1*×*10^−9^), are reported associations with haematological traits^63^. We identify two SNPs, rs11778922 and rs61955499, with female-specific effects on BMI change. rs11778922 (OR=0.984 for membership in the high-gain BMI cluster, *P* = 1.3 *×* 10^−8^, sex-heterogeneity *P*_*sex−het*_ = 5.8 *×* 10^−4^, see Methods) has previously been nominally associated with BMI in females^43^, and rs61955499 (OR=1.070 for membership in the BMI loss cluster, *P* = 3.4 *×* 10^−8^, *P*_*sex−het*_ = 4.7 *×* 10^−5^), has previously been nominally associated with low-density lipoprotein (LDL) cholesterol levels^64^. Finally, rs12953815 is associated with male-specific weight change (OR=1.012 for membership in the weight loss cluster, *P* = 1.7 *×* 10^−8^, *P*_*sexhet*_ = 2.0 *×* 10^−5^) and has been previously nominally associated with lung function^65^.

The smaller number of independent GWS associations with adiposity change: 6, compared to 374 unique lead SNPs associated with baseline obesity traits, which is expected given the 7- to 9-fold lower heritability of adiposity change. The heritability explained by genotyped SNPs 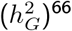 of the posterior probability of belonging to an adiposity-gain cluster is between 1.38% in men to 2.82% in women, while the 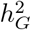 of baseline obesity traits varies between 21.6% to 29.0% across strata (**Figure 4**). Furthermore, we observe that the heritability of BMI and weight trajectories are higher in women than in men (2.89% vs 1.05% for BMI slopes, *P*_*sexhet*_ = 0.012; and 3.42% vs 1.69% for weight slopes, *P*_*sexhet*_ = 9.9 *×* 10^−3^). We do not observe a corresponding difference in the 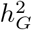 of baseline BMI or weight between the sexes (*P*_*sexhet*_ *>* 0.1). Finally, baseline and change in obesity traits are genetically correlated, with *r*_*G*_ ranging from 0.35 (95% CI=0.24-0.45) for weight in women to 0.91 (0.59-1.23) for BMI in men (**Figure 4**). While the genetic correlation between baseline adiposity and adiposity change appears to be higher in men as compared to women, these estimates have wide confidence intervals (overlapping 1) and *P*_*sexhet*_ *>* 0.05 for both BMI and weight.

**Figure 4:**
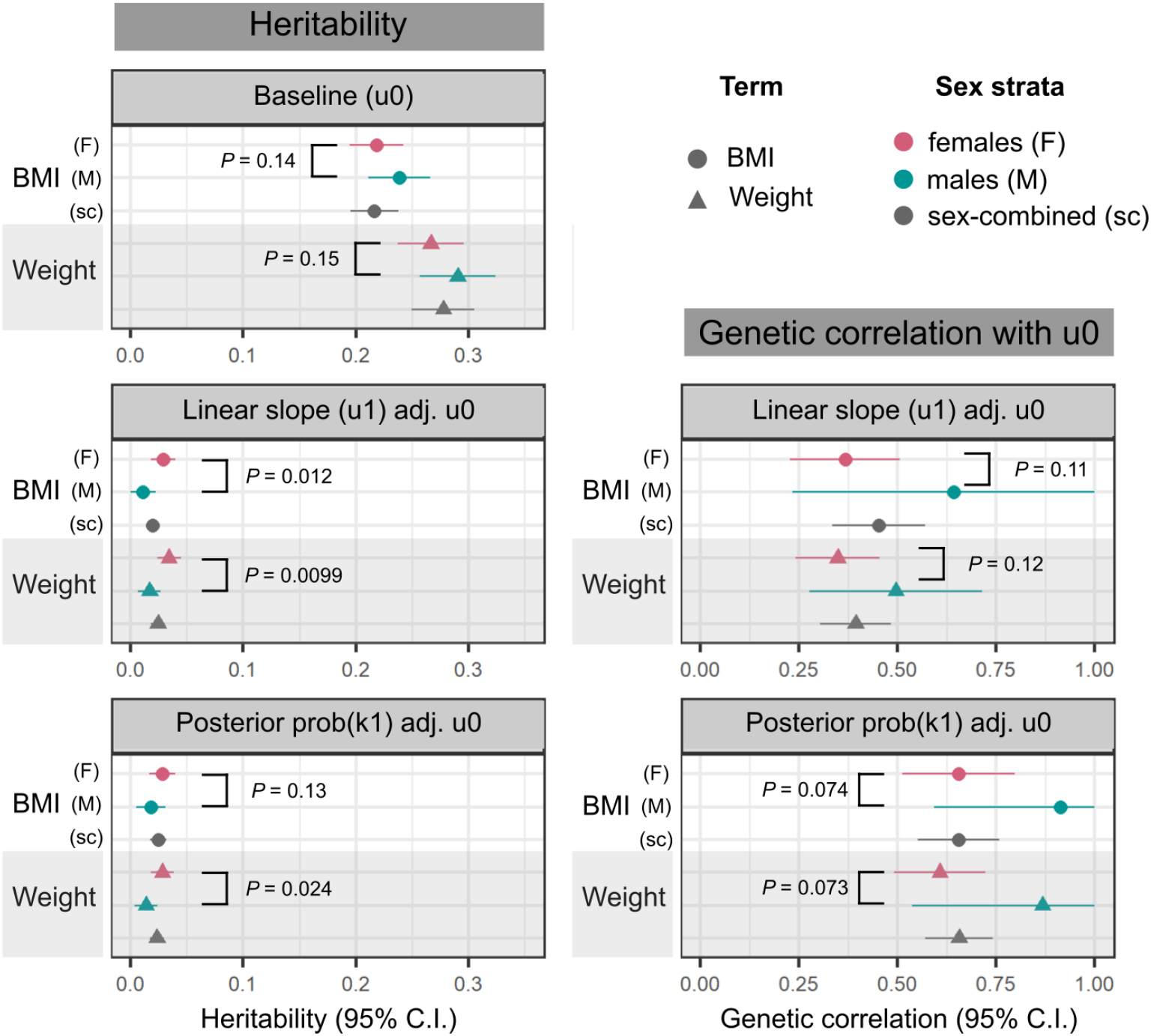
Genotyped SNP-based heritability of, and genetic correlation between, baseline obesity trait and obesity-change phenotypes. Left column: heritability 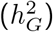 estimates and 95% CI, calculated using the LDSC software^66^ on a subset of 1 million HapMap3 SNPs^67^ for the following traits: baseline BMI and weight, estimated from intercepts of linear mixed-effects models of obesity traits over time (u0), linear slope change in obesity traits over time (u1 adj. u0), adjusted for intercepts, and posterior probability of membership in a high-gain BMI or weight cluster, adjusted for baseline trait value (prob(k1) adj. u0). Right column: Genetic correlation, *r*_*G*_ and 95% CI between the two obesity-change phenotypes and corresponding baseline obesity traits. In all panels, circles represent BMI, triangles represent weight; points are coloured by analysis strata (pink: female-sepcific, green: male-specific, grey: sex-combined). P-values display the level of significance of heterogeneity between the female- and male-specific estimates in each panel.

Throughout this study, we evaluate both BMI and weight as obesity traits, and expect these to track closely in adults as height does not change significantly over time. In the 161,891 individuals in our discovery strata with multiple measurements of both BMI and weight, there is a strong correlation between the slopes for weight and BMI change (*r*^2^ = 0.88) and between the posterior probabilities of membership in the BMI-gain and weight-gain clusters (*r*^2^ = 0.73) (**Supp. Table 9**, all *P <* 1 *×* 10^−16^). Moreover, the genetic correlation between change in BMI and weight is nearly perfect (*r*_*G*_ for slope terms=0.98, *r*_*G*_ for posterior probability of membership in gain cluster=0.95, all *P <* 1 *×* 10^−16^), indicating that the genetic architecture highlighted here is robust to the metric of adiposity used to define trajectories.

## Discussion

In this large-scale EHR- and genetics-based study of longitudinal trajectories of obesity traits, we demonstrate that modelling multiple observations across time increases power to identify genome-wide signals for baseline BMI and weight and enables the discovery of genetic variants associated with changes in adiposity, which are less heritable than and only partially shared with baseline adiposity. Modelling 1.5 million observations of BMI and weight from >177,000 individuals across time enabled us to identify 14 novel, biologically plausible, genetic signals associated with obesity traits. The discovery of these novel loci highlights that repeat measurements can contribute to narrowing the “missing heritability” gap. Leveraging the bespoke longitudinal adiposity phenotypes developed here, we find six genetic loci associated with changes in BMI and weight over time. While previous studies have investigated the associations of cross-sectional BMI SNPs or obesity polygenic scores with adiposity trajectories^15, 68^, to the best of our knowledge, this study reports the first genome-wide scan of variants associated with obesity trait trajectories over adulthood.

Accounting for the influence of genetic variation on adiposity change may provide opportunities to personalise obesity prevention and treatment^69, 70^. While several studies have investigated the association between BMIrelated genetic variants and weight loss guided by medical^71^, surgical^72, 73^, dietary^74^, or behavioural^71, 75–77^ interventions, results are inconsistent across studies, intervention types, and genes assessed. Given our evidence that the genetic basis of adiposity change is distinct from baseline levels, we hypothesise that genetic variants associated with longitudinal weight trajectories may be better predictors of long-term weight change following treatment or lifestyle interventions than variants associated with baseline BMI. Moreover, incorporating information on the genetic signals associated with adiposity trajectories will complement current genetics-based strategies to identify genes for pharmaceutical targets^78^ for obesity treatment.

Leveraging EHR to derive longitudinal metrics for genetic discovery may be affected by various biases described earlier^79^. However, the robustness of our results suggests that our discovery dataset may have mitigated against these biases in three ways: (1) While EHR data over-represent sick patients and individuals with higher BMI, UKBB participants are, on average, healthier and have lower BMIs than the population of the United Kingdom (UK)^80^. Therefore our UKBB-linked EHR discovery cohort is more overweight than a random sampling of UKBB, but in contrast UKBB itself is ascertained towards lower-BMI individuals than a random sampling of the UK. (2) Appending the more accurate UKBB assessment centre measurements to the EHR data improves overall data quality. (3) Stringent quality control at both the population and individual level reduces the signal to noise ratio by filtering out a subset of inaccurate data entries. Linking EHRs with biobank data may therefore provide a robust framework for genetic discovery.

The two-stage nature of our approach to associate genetic variants with longitudinal trajectories of obesity traits is highly advantageous because of its computational efficiency and convenience. In particular, our method is *composable*, as the longitudinal analysis of raw data can first be performed separately using a choice of popular, efficient implementations of models; the first-stage outputs can then be taken forward to a GWAS performed in its own bespoke, highly optimised software. The two-stage method approximates the fitting of a full joint model incorporating raw measurement data and genome-wide SNP data. While a full joint model would propagate posterior uncertainty from the longitudinal sub-model through to the GWAS, the approximation here takes forward a single point estimate, i.e. a best linear unbiased predictor (BLUP) or posterior probability of cluster membership, to GWAS. However, in EHR datasets, the number of measurements, and hence estimation precision, can vary across individuals. Hence, the propagation of uncertainty between model components, in a similar vein to Markov melding^81^, has the potential to further increase power for genetic discovery. An interesting area for future research will be to allow for the principled propagation of posterior uncertainty in inputted traits through to the highly optimised, multi-locus, mixed-model GWAS methods to perform genetic association in the presence of relatedness and population stratification^82^.

It is also important that the choice of trajectory metric utilised in genetic analysis is phenotype-aware. While the variance within an individual’s trait value over time may capture meaningful biology for biomarkers such as blood pressure or triglycerides, whose fluctuations are associated with disease development and progress^83, 84^, weight is a more stable trait which shows a steady pattern of change over many years^85, 86^. Our adiposity-change metrics, derived from regression models incorporating linear and non-linear temporal trends, are better suited to identify the genetic component of BMI and weight trajectories, and are robust to the manner in which this is defined. For example, many of the lead SNPs from our obesity-change GWASs are also associated with self-reported weight change, despite self-report being an imprecise metric^87^.

In particular, rs429358 (missense variant in *APOE*) is robustly associated with loss in BMI and weight, independent of baseline obesity, across men and women, in individuals of various ancestral groups. *APOE* codes for apolipoprotein E, which is a core component of plasma lipoproteins that is essential for cholesterol transport and homeostasis in several tissues across the body, including the central nervous system, muscle, heart, liver, and adipose tissue^88, 89^. The precise pathway by which this variant affects weight change is difficult to pinpoint, as *APOE* is a highly pleiotropic locus associated with hundreds of biomarkers and diseases^62^. Here too, we find an association between rs429358 and changes in 11 biomarkers over time. Obesity is cross-sectionally associated with several of these, including positive correlations with levels of triglycerides and total cholesterol^90, 91^, markers of chronic inflammation^92^, and haematological traits^93^, and negatively correlated with levels of HDL cholesterol^90, 91^ and potassium^94^; however, the longitudinal and causal nature of these associations remain to be established. As rs429358 is also the strongest genetic risk factor for Alzheimer’s disease^58, 59^, which is preceded by weight loss^95^, we ensured that our findings were robust to the exclusion of individuals with dementia. We hypothesise that the *APOE* effect on weight loss may act through cholesterol- and lipid-metabolism pathways that partly determine response to dietary and environmental factors, as seen in mouse models^96, 97^. Indeed, it has recently been suggested that *APOE* -mediated cholesterol dysregulation in the brain may influence the onset and severity of Alzheimer’s disease^98^, suggesting that ageing-associated systemic aberrations in cholesterol homeostasis could have far-ranging consequences from weight loss to cognitive decline.

Patterns of weight change in mid-to-late adulthood have been observed to be sex-specific, particularly as women undergo significant changes in weight and body fat distribution around menopause^99^. Here, we find that the heritability of changes in obesity traits is significantly higher in women than in men, supporting a previous finding that obesity polygenic scores are more strongly associated with weight-change trajectories in women than in men^68^. This is in contrast to baseline obesity, which is equally heritable in men and women, both in our study and as previously reported^43^. The lower genetic correlation between baseline obesity and obesity-change in women as compared to men, while not statistically significant, may nevertheless indicate sex-differential genome-wide contributions to these phenotypes. We hypothesise that sex hormones could explain some of this sex-specificity, particularly through their role in altering overall obesity and fat distribution around menopause^100, 101^. We were under-powered to study the genome-wide architecture of change in adult WC and WHR (ten-fold fewer observations than BMI and weight), whose cross-sectional levels are genetically sex-specific with higher heritability in women^43^, so more work is needed to disentangle the genetic contribution to changes in adult body fat distribution over time.

While the EHR-linked UKBB cohort has driven genetic discovery for a vast array of human traits in populations of European ancestry^102^, sample sizes remain under-powered to detect genome-wide associations in other ancestral groups. We were thus limited to replicating European-ancestry associations in other populations, without the ability to discover ancestry-specific variants associated with adult adiposity trajectories. Furthermore, despite the inclusion of >200,000 individuals in the first release of the UKBB EHR data, sample sizes remain low to analyse the genetics of longitudinal trajectory metrics, which have lower heritability than the averaged trait value^15, 103^ (*^*7-9x lower in our study) and are thus more challenging to characterise genetically without corresponding increases in sample size. Another limitation of our study was the exclusion of time-varying covariates, such as medication use, smoking status, and other dietary and environmental covariates from models of adiposity change. It is challenging to extract time-dependent values of these variables from EHRs and difficult to ascertain the direction of causality by which these covariates may be associated with weight change. For example, the use of statins to lower blood pressure may be connected to weight gain, mediated indirectly by change in appetite^104^, but high blood pressure may itself be a consequence of weight gain^105^. Inappropriate adjustments along this causal pathway may lead to unexpected collider biases^106^. In general, despite their longitudinal nature, it is challenging to assign causality to the associations between weight change and covariates or disease diagnoses from EHR observations alone, as there is no prospective study design to follow^107^. Advances in emulating randomised control trials from longitudinal EHR are beginning to overcome these challenges^108, 109^, and in the future, it will be critical to incorporate information on genetic risk into these simulated studies.

To the best of our knowledge, this is the largest study to date that characterises the genome-wide architecture of adult adiposity trajectories, and the first to identify specific variants that alter BMI and weight in mid- to late-adulthood. We add evidence to support the growing utility of EHRs in genetics research, and particularly highlight opportunities for incorporating longitudinal information to boost power and identify novel associations. In particular, the *APOE* -associated weight loss identified here contributes to a growing body of evidence on the ageing-associated effects of cholesterol dysregulation. Heterogeneity between men and women in the genome-wide architecture of obesity-change and genetic correlation with baseline obesity highlights the importance of distinguishing between the genetic contributions to mean and lifetime trajectories of phenotypes in sex-specific analyses. In the future, the growing integration of EHR with genetic data in large biobanks will allow us to assess the time-varying associations of rare variants with outsize effects on quantitative traits, as well as to establish genetic and phenotypic relationships among the trajectories of multiple correlated biomarkers across adulthood.

## Methods

### Identification and quality control of longitudinal obesity records

#### UK Biobank

This study was conducted using the UKBB resource, which is a prospective UK-based cohort study with approximately 500,000 participants aged 40–69 years at recruitment, on whom a range of medical, environmental, and genetic information has been collected^42^. Here, we included 409,595 individuals in the white British ancestry subset identified by Bycroft *et al*.^110^ who passed genotype quality control (QC) (see below).

#### Repeat obesity trait measurements

Obesity-associated traits including BMI and weight were recorded at initial baseline assessment (between 2006-2010), as well as at repeat assessments of 20,345 participants (between 2012-13), and at imaging assessments of 52,596 participants (in 2014 and later). We curated a longitudinal research resource by integrating these repeat UKBB assessment centre measurements with the interim release of primary care records provided by GPs for approximately 45% of the UKBB cohort (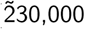 participants, randomly selected)^111^ (**Supp. Fig. 3**). Each individual with at least one BMI record (coded as Clinical Practice Research Datalink (CPRD) code 22K..) or weight record (coded as CPRD code 22A..) in the GP data had their respective UKBB assessment centre measurements appended. Following phenotype and genotype QC, this resulted in 162,666 participants of white British ancestry with multiple BMI measurements and 177,472 participants with multiple weight measurements (**Supp. Fig. 3**).

#### Quality control

We performed both population-level and individual-level longitudinal QC. Participants with codes for history of bariatric surgery (**Supp. Table 10**, as identified by Kuan *et al*.^112^) were excluded entirely, while BMI and weight observations up to the date of surgery were retained for individuals where this could be determined. Only those measures recorded in adulthood (ages 20 - 80 years) were retained. We excluded implausible observations, defined as more extreme than +/- 10% of the UKBB asessment centre minimum and maximum values respectively (BMI < 10.9 kg/m^2^ or > 82.1 kg/m^2^ and weight < 27 kg or > 217 kg). We further removed any extreme values > 5 SDs away from the population mean to exclude possible technical errors. At the individual level we excluded multiple observations on the same day, which are likely to be recording errors, by only retaining the observation closest to the individual’s median value of the trait across all time-points. Finally, we excluded any extreme measurements on the individual level. For individual *i* with *J*_*i*_ data points represented as (measurement, age) pairs (*y*_*i,j*_, *t*_*i,j*_) for *j* = 1, …, *J*_*i*_ ordered chronologically, i.e. *t*_*i*,1_ *<* … *< t*_*i,J*_*i*, a “jump” *P*_*i,j*_ for *j* = 1, …, *J*_*i*_ − 1 was defined as:

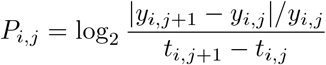

We removed data points associated with extreme jumps (> 3 SDs away from the population mean jump, to exclude possible technical errors) by excluding the observation farther from the individual’s median value of the trait across all time-points.

#### BMI and weight validation data

Participants with BMI and weight observations in UKBB assessment centre measurements who were not included in the interim release of the GP data were held out of discovery analyses (**Supp. Fig. 3**). This resulted in 245,447 individuals with at least one BMI observation and 230,861 individuals with at least one weight observation for replication of cross-sectional results. For the replication of longitudinal results, a subset of individuals was used comprising 17,006 individuals with multiple observations of BMI, and 17,035 individuals with multiple observations of weight, from repeat assessment centre visits.

#### Self-reported weight change data

At each UKBB assessment centre visit, participants were asked the question: “Compared with one year ago, has your weight changed?”, reported as “No - weigh about the same”, “Yes - gained weight”, “Yes - lost weight”, “Do not know”, or “Prefer not to answer”. We coded the 1-yr self-reported weight change response at the first assessment centre visit as an ordinal categorical variable with three levels: “loss”, “no change”, and “gain”, excluding individuals who did not respond or responded with “Do not know” or “Prefer not to answer”. We retained 301,943 individuals of white British ancestry that were not included in any of the discovery analyses.

#### Abdominal adiposity data

Similar to the BMI and weight validation datasets, we retained the 44,154 participants with multiple WC and hip circumference (HC) records across repeat assessment centre visits who were not included in the interim release of the GP data, and hence held out of discovery analyses. WHR was calculated at each visit by taking the ratio of WC to HC. We further calculated WC adjusted for BMI (WCadjBMI) and WHR adjusted for BMI (WHRadjBMI) values at each visit for which WC, HC, and BMI were recorded simultaneously by taking the residual of WC and WHR in linear regression models with BMI as the sole predictor.

### Models to define baseline adiposity and adiposity change traits

Individual *i* has *J*_*i*_ data points represented as (measurement, age) pairs (*y*_*i,j*_, *t*_*i,j*_) for *j* = 1, …, *J*_*i*_ ordered chronologically, i.e. *t*_*i*,1_ *<* … *< t*_*i,J*_*i*. The following models are all fitted separately in three strata: female-specific, male-specific, and sex-combined.

#### Intercept and slope traits for GWAS

We implement a two-stage algorithm to estimate and preprocess local intercept and slopes of obesity traits to be taken forward to GWAS in both discovery and validation datasets.

1. Fit random-slope, random-intercept mixed model with the maximum likelihood estimation procedure in the `lme4`^113^ package in `R`^114^. We target two quantities: the baseline value of each individual’s clinical trait (the *β*_0_ + *u*_*i*,0_ below); and the the linearly approximated rate of change in the trait during each individual’s measurement window (the *β*_1_ + *u*_*i*,1_ below):

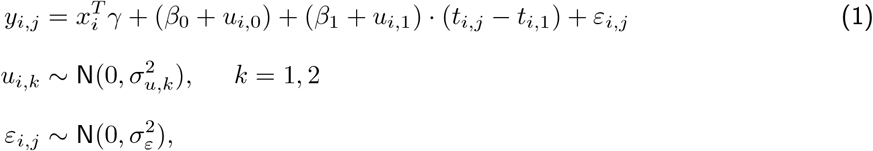

where individual-specific covariates *x*_*i*_ comprise: baseline age, (baseline age)^2^, data provider, year of birth, and sex. Variance parameters 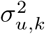 and 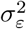 are estimated. Fitting model (1) outputs fixed effect model estimates 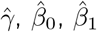 and BLUPs of the random effects 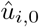 and 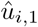.
2. Linearly adjust and transform the outputted BLUPs. We fit and subtract the linear predictor in each of the linear models:

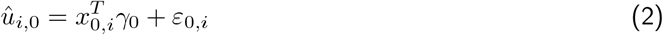

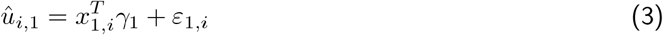

where the intercept-adjusting covariates *x*_0,*i*_ in (2) comprise: baseline age, (baseline age)^2^, sex, year of birth, assessment centre, number of follow-ups, and total length of follow-up (in years). Slope-adjusting covariates *x*_1,*i*_ in (3) comprise the same as *x*_0,*i*_ but additionally include the intercept BLUP *û*_*i*,0_. We finally apply a deterministic rank-based inverse normal transformation^115^ to the residuals from fitting models (2) and (3). For example, the intercept trait for individual *i* taken forward to GWAS is

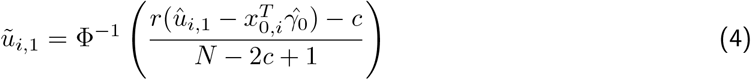

where 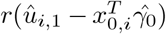 is the rank of the *i*th residual among all *N* residuals, the offset *c* is 0.5, and Φ(*·*) is the cumulative distribution function (CDF) of the standard Gaussian distribution.

#### Modelling nonlinear trajectories with regularised splines

We model non-linear changes in obesity traits using a regularised B-spline basis of degree 3 (i.e., a cubic spline model) with *n*_df_ = 100 degrees of freedom, incorporating *n*_df_ − 4 (i.e., *n*_df_ − 3 [degree] − 1 [intercept]) knots that are spaced evenly across each individual’s first *T* = 7500 post-baseline days *≈* 20.5 years. It is common practice in semi-parametric regression to use *regularised* splines with a relatively *large* number of knots, thereby allowing functional expressiveness without overfitting^31, 116^. Conditional on the spline coefficients, ***b***_*i*_, the likelihood for measurements ***y***_*i*_ (individual *i*’s *J*_*i*_-vector of measurements taken at days 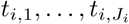) is

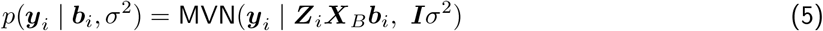

where: the *n*_df_-vector ***b***_*i*_ contains the *i*th individual’s spline basis coefficients; ***X***_*B*_ is the (*T* + 1) *× n*_df_ matrix of spline basis functions evaluated at days 0, …, *T* post-baseline; and ***Z***_*i*_ is a *J*_*i*_ *×* (*T* + 1) matrix whose *j*th row extracts day *t*_*i,j*_ *− t*_*i*,1_ post-baseline, i.e.,

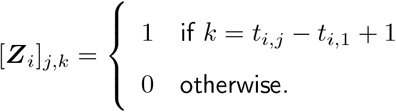

We specify an order-1 autoregressive (*AR*(1)) model as a smoothing prior on spline coefficients, ***b***_*i*_, which vary smoothly around an individual-specific mean value, *µ*_*i*_. On *µ*_*i*_ we specify a non-informative prior: 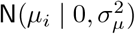 with large SD *σ*_*µ*_. The resulting *µ*_*i*_-marginalised prior for ***b***_*i*_ is

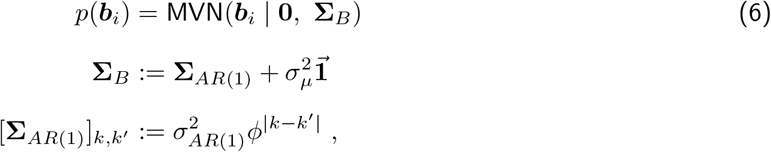

where: **Σ**_*AR*(1)_ is the *n*_df_ *× n*_df_ autocovariance matrix implied by an *AR*(1) model with lag-1 autocorrelation *ϕ ∈* [0, 1) and scale parameter 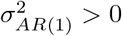; and 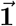is an *n*_df_ *× n*_df_ matrix of ones.

The prior at (6) and likelihood at (5) are a specific case of the Bayes linear model^117^, for which the posterior is available in closed form:

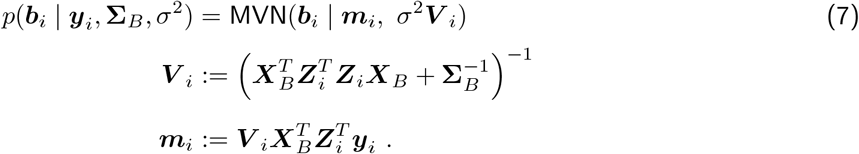

The posterior at (7) can be evaluated separately and in parallel across individuals because the (***y***_*i*_, ***b***_*i*_) are conditionally independent across individuals *i* given the hyperparameters 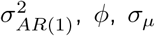 and *σ*^2^. Values of hyperparameters in the smoothing prior are chosen subjectively, via visualisation of randomly selected samples of individual data trajectories, to reflect empirical levels of smoothness: 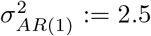, *ϕ*:= 0.99, *σ*_*µ*_ := 100 (**Supp. Fig. 4**). We additionally compared cluster allocations for 5,000 randomly selected individuals across the following settings of hyperparameters: (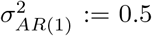, *ϕ* := 0.9, *σ*_*µ*_ := 10), (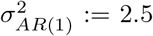, *ϕ* := 0.99, *σ*_*µ*_ := 100), and (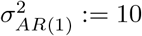, *ϕ* := 0.999, *σ*_*µ*_ := 500) (**Supp. Fig. 8**).

For each trait separately, we set *σ*^2^ to the median of its individual-specific maximum likelihood estimates (MLEs), i.e., 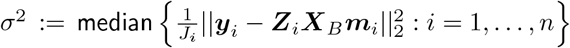 where each MLE is calculated from (5) after substituting for ***b***_*i*_ its maximum a posteriori estimate, ***m***_*i*_ from (7) (**Supp. Table 12**).

The measurements ***y***_*i*_ inputted into the likelihood for the regularised spline model at (5) are pre-processed by taking the standardised residual from the linear model with the following covariates: baseline age, (baseline age)2, data provider, year of birth, and sex, i.e. from the model 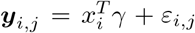 fitted across all *i* = 1, …, *N* individuals and *j* = 1, …, *J*_*i*_ time points. Standardisation of residuals then proceeds by subtracting the mean and dividing by the SD of residuals across all individuals and time points.

We focus on individual *i*’s posterior change from baseline, i.e. on

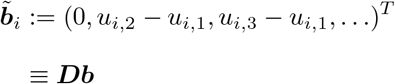

where the *j*th row of ***D*** is (***e***_*j*_ − ***e***_1_)^*T*^ and ***e***_*k*_ is the *k*th basis vector, i.e. a column *n*_df_-vector with zeroes everywhere except the *k*th entry, which is one. To calculate the posterior for 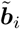 we linearly transform the posterior at (7) so that

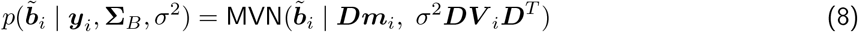

with ***m***_*i*_ and ***V*** _*i*_ defined at at (7).

#### Soft clustering of individuals by non-linear adiposity trajectory patterns

See **Supp. Fig. 5** for an overview of the clustering protocol.

Any two individuals typically have quite distinct measurement profiles, with different numbers of measurements taken at ages which may be quite disparate. Therefore the precision with which we can estimate any particular spline coefficient varies across individuals. To incorporate this heteroscedasticity into our clustering framework, we define the following scaled Euclidean distance between each pair of individuals (*i, i*^*′*^) in the space of baselined spline basis coefficients:

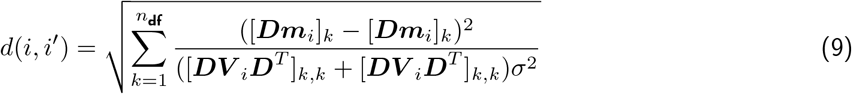

where ***m***_*i*_ and *σ*^2^***V*** _*i*_ are the posterior mean and covariance of individual *i*’s spine coefficients ***b***_*i*_ taken from (7). For each spline coefficient *k* in (9), the squared difference between individuals’ *i* and *i*^*′*^ mean coefficients is standardised by the sum of the corresponding variances.

We perform *k*-medoids clustering using the the Partitioning Around Medoids (PAM) algorithm^54, 55^ as implemented in the `pam` function in the `cluster` package^118^ in `R`^114^. We train cluster centroids on a randomly selected subset of 80% of individuals in each analysis strata. We filter individuals in the training set to retain only those with at least *L* = 2 observations. For a fixed number of clusters, *K* = 4, we initialize cluster membership according to bins *B*_1:*K*_ demarcated by the 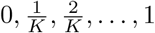 empirical quantiles of the estimated fold change in obesity trait between baseline and year *M* = 2:

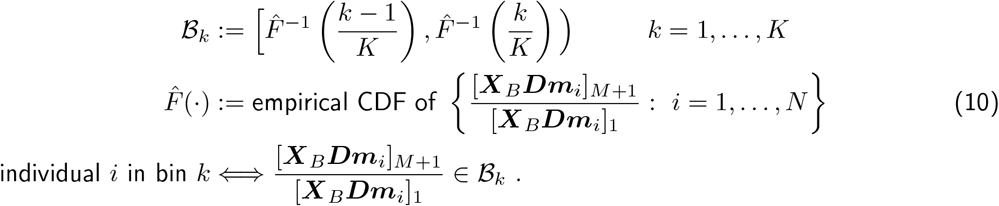

To ensure robustness, we run the clustering algorithm *S* = 10 times, each on a random sub-sample of size 5,000 (without replacement). For each clustering output *s* = 1, …, *S*, we calculate the point-wise mean of each clusters’ constituent individuals:

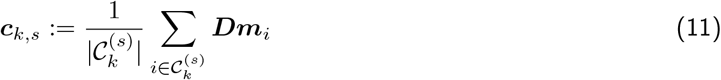

For each clustering *s*, we observe all trajectories ***c***_*s*,1:*K*_ to be monotonic and non-overlapping (**Supp. Fig. 6**). We can therefore define ordered cluster means ***c***_(*k*),*s*_,

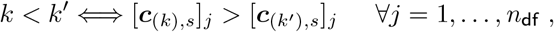

and average the *k*th ordered mean across *S* clusterings, where the highest-weight cluster mean is given by ***c***_(1)_ and the lowest by ***c***_(*K*)_:

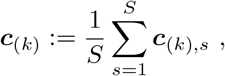

with corresponding point-wise standard errors (SEs). We investigate the sensitivity of the resulting clusters to number of clusters *K*, filter parameter *L* (minimum number of measurements), and the cluster initialisation parameter *M* appearing in (10) via silhouette values^119^, which evaluate the similarity between cluster members (cohesion) vs others (separation) (**Supp. Fig. 6**). We test values of *K* from 2, …, 8, filtering parameter *L ∈* (2, 5, 10), and initialisation parameter *M ∈* (1, 2, 5, 10) or random initialisation to choose a combination of parameters that produces dense and separable clusters, i.e. *K* = 4, *L* = 2, *M* = 2. We also qualitatively evaluate cluster centroids across all parameter settings (**Supp. Fig. 7**). Finally, we compared cluster allocations over each of the 10 random trains for a set of 5,000 randomly sampled individuals held out of the training splits (**Supp. Fig. 9**).

Once cluster centroids have been calculated, we define individual *i*’s soft cluster membership probability of belonging to cluster *k* as the posterior probability of being closest in Euclidean distance to cluster *k*’s centroid:

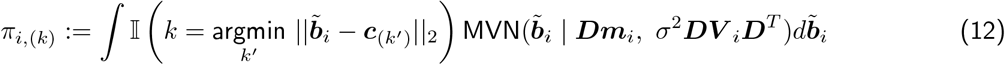

where the second term in the integrand is the posterior from (7), and we approximate the integral in (12) using 100 Monte Carlo samples from the posterior.

Finally, we validate the clustering by comparing cluster properties of the randomly selected 80% training set used to define cluster centroids, with the held-out 20% validation set. We assign each individual to the cluster for which they have highest membership probability and compare the proportion of individuals assigned to each cluster, as well as distributions of sex, baseline age, number of follow-up measures, and total length of follow-up of individuals assigned to each cluster. These metrics are similar across training and validation sets in all strata (**Supp. Table 13**).

Finally we take forward bounded logit-transformed cumulative cluster probabilities to GWAS. These outputs are defined as bounded logit(*π*_*i*,(1)_), bounded logit(*π*_*i*,(1)_ + *π*_*i*,(2)_), and bounded logit(*π*_*i*,(1)_ + *π*_*i*,(2)_ + *π*_*i*,(3)_), i.e., the bounded log odds of being in the highest (k1), highest two (k1 or k2), and highest three (k1, k2 or k3) weight clusters respectively. To prevent infinite log odds at *π ∈ {*0, 1*}* we defined the following bounded logit transform^120^:

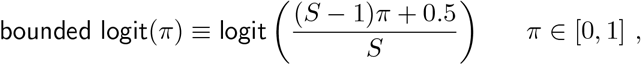

where *S* = 100, the number of Monte Carlo samples from the posterior in approximating (12).

### Genome-wide association studies

#### QC of UK Biobank genotyped and imputed data

Genotyping, initial genotype QC, and imputation were performed by UKBB^110^. We performed post-imputation QC to retain only bi-allelic SNPs with MAF > 0.01, info score > 0.8, missing call rate < 5%, and Hardy-Weinberg equilibrium (HWE) exact test *P >* 1 *×* 10^−6^. We additionally performed sample QC to exclude individuals with sex chromosome aneuploidies, whose self-reported sex did not match inferred genetic sex, with an excess of third degree relatives in UKBB, identified as heterozygosity or missingness outliers, excluded from autosome phasing or kinship inference, and any other UKBB recommended exclusions^110^.

#### Linear mixed model association analyses for quantitative traits

The following association analyses are all performed separately in three strata: female-specific, male-specific, and sex-combined. The intercept and slope traits for GWAS, i.e. 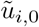 and 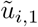 were tested for association with genetic variants, adjusted for the first 21 genetic principal components (PCs) and genotyping array, using the `BOLT-LMM` software^82^. A similar protocol was followed for the logit-transformed soft clustering probability traits, i.e. 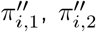, and 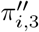 with additional adjustments for baseline trait, baseline age, (baseline age)^2^, sex, year of birth, assessment centre, number of follow-ups, and total length of follow-up (in years).

#### Fine-mapping SNP associations

We identified putative causal variants at all GWS loci (defined by merging windows of 1.5 Mb around SNPs with *P <* 5 *×* 10^−8^), using FINEMAP^121^ to select variants (lead SNPs) with a posterior inclusion probability > 95%. Lead SNPs were annotated to the nearest gene transcription start site.

#### Classifying baseline BMI and weight SNPs as reported, refined, or novel obesity associations

We curated a list of SNPs associated with any of 44 obesity-related traits in the GWAS Catalog^53^ accessed on 02 Nov 2021, henceforth referred to as “published obesity-associated variants” (**Supp. Table 1**). We then conducted conditional analysis using `GCTA-COJO`^122^ for each lead SNP in our GWAS and published obesity-associated variants within 500 kb, classifying variants as reported, refined, or novel based on previously recommended criteria^44^. Reported SNPs in our study are those whose effects are fully accounted for by published obesity-associated variants within 500 kb. Refined SNPs fulfill all of the following criteria: (1) the refined SNP is correlated (LD *r*^2^ *≥* 0.1) with at least one published obesity-associated variant within 500 kb, (2) the refined SNP has a significantly stronger effect (*P <* 0.05 in a two-sample t-test for difference in mean effect sizes) on the BMI- or weight-intercept trait than published obesity-associated SNPs and also accounts for the effect of published obesity-associated SNPs in conditional analysis (conditional *P >* 0.05), and (3) published obesity-associated SNPs cannot fully account for the effect of the refined SNP in conditional analysis (conditional *P <* 0.05). Finally, a SNP in our study was declared novel if it was not in LD with (*r*^2^ < 0.1), and conditionally independent of (conditional *P <* 0.05), all published obesity-associated variants within 500 kb.

### Replication of GWS associations in UK Biobank hold-out sets

#### BMI and weight intercept-trait genetic associations

We created cross-sectional obesity phenotypes for the 245,447 individuals in the hold-out set for BMI and 230,861 individuals in the hold-out set for weight (**Supp. Fig. 3**) by retaining the observed trait value closest to the individual’s median trait value (if multiple observations present). Deterministic rank-based inverse normal transformation^115^ was applied to the residual of the obesity trait adjusted for age, age^2^, year of birth, data provider, and sex. We then tested this trait for association with genetic variants, adjusted for the first 21 genetic PCs and genotyping array, using the `BOLT-LMM` software^82^.

#### BMI and weight slope-trait genetic associations

We created adiposity slope phenotypes for the 17,006 individuals with multiple observations of BMI and 17,035 individuals with multiple observations of weight from repeat assessment centre visits (**Supp. Fig. 3**) with BLUPs from linear mixed-effects models as described in the slope trait modelling section above. We tested for association of this slope trait with GWS variants associated with adiposity change in our discovery analyses, adjusted for the first 21 genetic PCs and genotyping array, via the linear regression framework implemented in `PLINK`^123^. As `PLINK` does not account for family structure, we compared each pair of second-degree or closer related individuals (kinship coefficient > 0.0884)^110^ and excluded the individual in the pair having higher genotyping missingness. We repeated the same protocol within each self-identified ethnic group of individuals not of white British ancestry.

#### Genetic associations with BMI and weight cluster probabilities

We fit regularised splines as detailed above to the 17,006 individuals with multiple observations of BMI and 17,035 individuals with multiple observations of weight from repeat assessment centre visits (**Supp. Fig. 3**). Soft cluster membership probabilities for these individuals were calculated, and the three logit-transformed *π*_*i*_ traits were carried forward for association testing with GWS variants associated with adiposity change in our discovery analyses. As above, we pruned out second-degree or closer related individuals and performed association analysis, adjusted for baseline trait, baseline age, (baseline age)^2^, assessment centre, first 21 genetic PCs and genotyping array, via the linear regression framework implemented in `PLINK`^123^. We repeated the same protocol within each self-identified ethnic group of individuals not of white British ancestry.

#### Genetic associations with self-reported weight change

We fit proportional odds logistic regression models implemented in the `MASS` package^124^ in `R`^114^ to estimate the additive effect of lead SNPs on self-reported one-year weight change coded as an ordinal categorical variable with three levels: “loss”, “no change”, and “gain” in 301,943 individuals (described in the data section above). All models were adjusted for BMI, age, sex, year of birth, data provider, assessment centre, first 21 genetic PCs and genotyping array. We repeated the same protocol within each self-identified ethnic group of individuals not of white British ancestry.

### Power comparison to GIANT 2019 meta-analysis of BMI

We accessed publicly available summary statistics from the GIANT consortium’s meta-analysis of BMI across UKBB and previous GIANT releases in female-specific (max N=434,793), male-specific (max N=374,755), and sex-combined strata (max N=806,834)^43^. SNPs included in both the GIANT 2019 meta-analysis and our in-house BMI-intercept GWAS that reached GWS in either study were carried forward for power comparisons, resulting in 26,812 (female-specific strata), 22,123 (male-specific strata), and 82,559 (sex-combined strata) SNPs. Per variant, we calculated the *χ*^2^ statistic (as 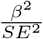) and obtained the ratio of 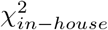 to 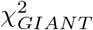. Median 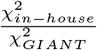 across all GWS SNPs was then compared to the median ratio of sample sizes, i.e. 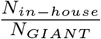,to determine the boost in power over that expected from the sample size difference between the two studies.

### rs429358 single-variant analyses

The following analyses were all conducted in female-specific, male-specific, and sex-combined strata.

#### Abdominal adiposity change traits

Slope changes in WC, WHR, WCadjBMI, and WHRadjBMI for up to 44,154 individuals with repeat observations were calculated using linear mixed-effects models, adjusted and rank-based inverse-normal transformed^115^ for genetic association testing as described in the slope modelling section above. We estimated the additive association of number of copies of the rs429358 minor allele (0, 1, or 2) with slope traits adjusted for the first 21 genetic PCs and genotyping array via linear regression.

#### Longitudinal phenome-wide association

We curated a longitudinal research resource for 45 additional quantitative phenotypes in up to 146,099 individuals of white British ancestry (**Supp. Table 14**, as identified by Kuan *et al*.^125^) by integrating UKBB assessment centre measurements with the interim release of primary care records provided by GPs, with QC performed as described above for obesity traits. Slope changes in each of these phenotypes were calculated using linear mixed-effects models described in (1). A deterministic rank-based inverse normal transformation^115^, as described in (4), was applied to the slope BLUP *û*_*i*,1_. The transformed slope trait was tested for additive association with number of copies of the rs429358 minor allele (0, 1, or 2), adjusted for the intercept BLUP *û*_*i*,0_, baseline age, (baseline age)2, sex, year of birth, number of follow-ups, total length of follow-up (in years), assessment centre, first 21 genetic PCs and genotyping array.

#### Identification of individuals with Alzheimer’s or dementia diagnoses

We identified participants with codes for history or diagnosis of dementia in either primary care or hospital in-patient records (**Supp. Table 15**, as identified by Kuan *et al*.^112^). We performed sensitivity analyses for the replication of rs429358 associations with all obesity-change phenotypes after excluding up to 242 individuals of white British ancestry with recorded history or diagnosis of dementia.

### SNP heritability and genetic correlations

We estimated the heritability explained by genotyped SNPs 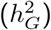 and genetic correlations (*r*_*G*_) between obesity-intercept and obesity-change traits, from summary statistics, using LD score regression implemented in the *LDSC* software^66, 126^, with pre-computed LD-scores based on European-ancestry samples of the 1000 Genomes Project^127^ restricted to HapMap3 SNPs^67^. The same protocol was followed to determine *r*_*G*_ between BMI-intercept in our in-house study and BMI in the GIANT 2019 meta-analysis.

### Sex heterogeneity testing

We tested for sex heterogeneity in the effects of adiposity-change lead SNPs by calculating Z-statistics and corresponding P-values for the difference in female-specific and male-specific effects as:

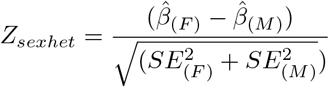

A similar statistic and test was used to determine heterogeneity between 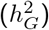 of all traits in males and females, and *r*_*G*_ between obesity-intercepts and obesity-change traits in males and females.

## Data Availability

All generated summary statistics from GWAS will be made publicly available through the GWAS Catalog upon publication. All code required to reproduce analyses will be made publicly available at: https://github.com/lindgrengroup/longitudinal_primarycare/tree/main/adiposity/scripts/manuscript.

## Data and code availability

All generated summary statistics from GWAS will be made publicly available through the GWAS Catalog^53^ upon publication. All code required to reproduce analyses are publicly available at: *https://github.com/lindgrengroup/longitudinal_primarycare/tree/main/adiposity/scripts/manuscript*.

## Acknowledgements

S.S.V. is supported by the Rhodes Scholarship, Clarendon Fund, and the Medical Sciences Doctoral Training Centre at the University of Oxford. K.C. is supported by the University of Leicester (College of Life Sciences) and Health Data Research UK. L.B.L.W. is supported by the Wellcome Trust (221651/Z/20/Z). C.H. is supported by the Alan Turing Institute, the EPSRC grant Bayes4Health, Novartis, and Novo Nordisk. C.M.L. is supported by the Li Ka Shing Foundation, NIHR Oxford Biomedical Research Centre, Oxford, NIH (1P50HD104224-01), Gates Foundation (INV-024200), and a Wellcome Trust Investigator Award (221782/Z/20/Z). The research was supported by the Wellcome Trust Core Award Grant Number 203141/Z/16/Z with additional support from the NIHR Oxford BRC. This research has been conducted using the UK Biobank Resource under Application Number 10844. The views expressed are those of the authors and not necessarily those of the NHS, the NIHR or the Department of Health.

