## Supplementary figures and images for "The genetic architecture of changes in adiposity during adulthood"

### Supp-Fig-1-supp-results-nonwb-ancestry.png

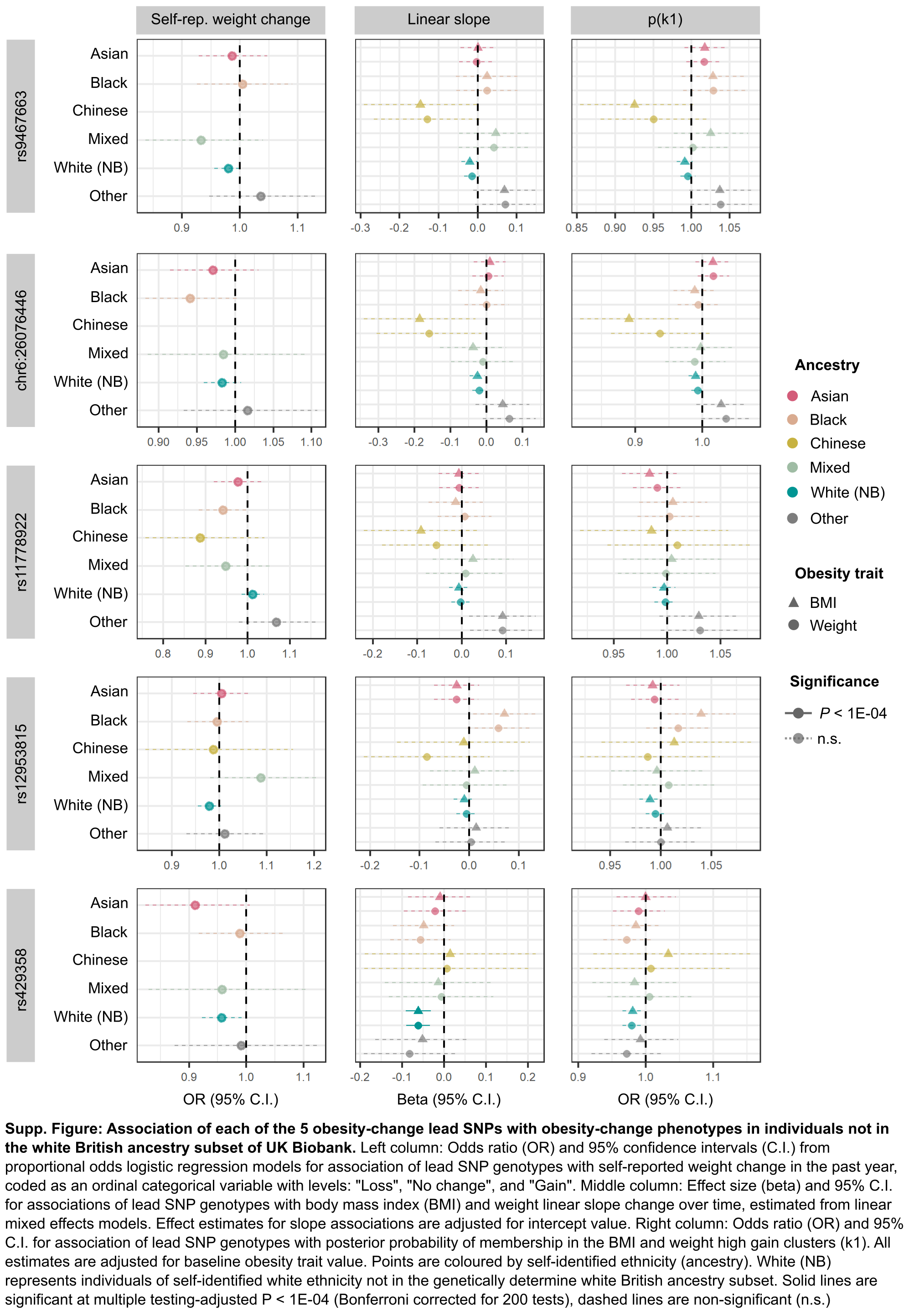

### Supp-Fig-2-supp-results-rs429358-dementia.png

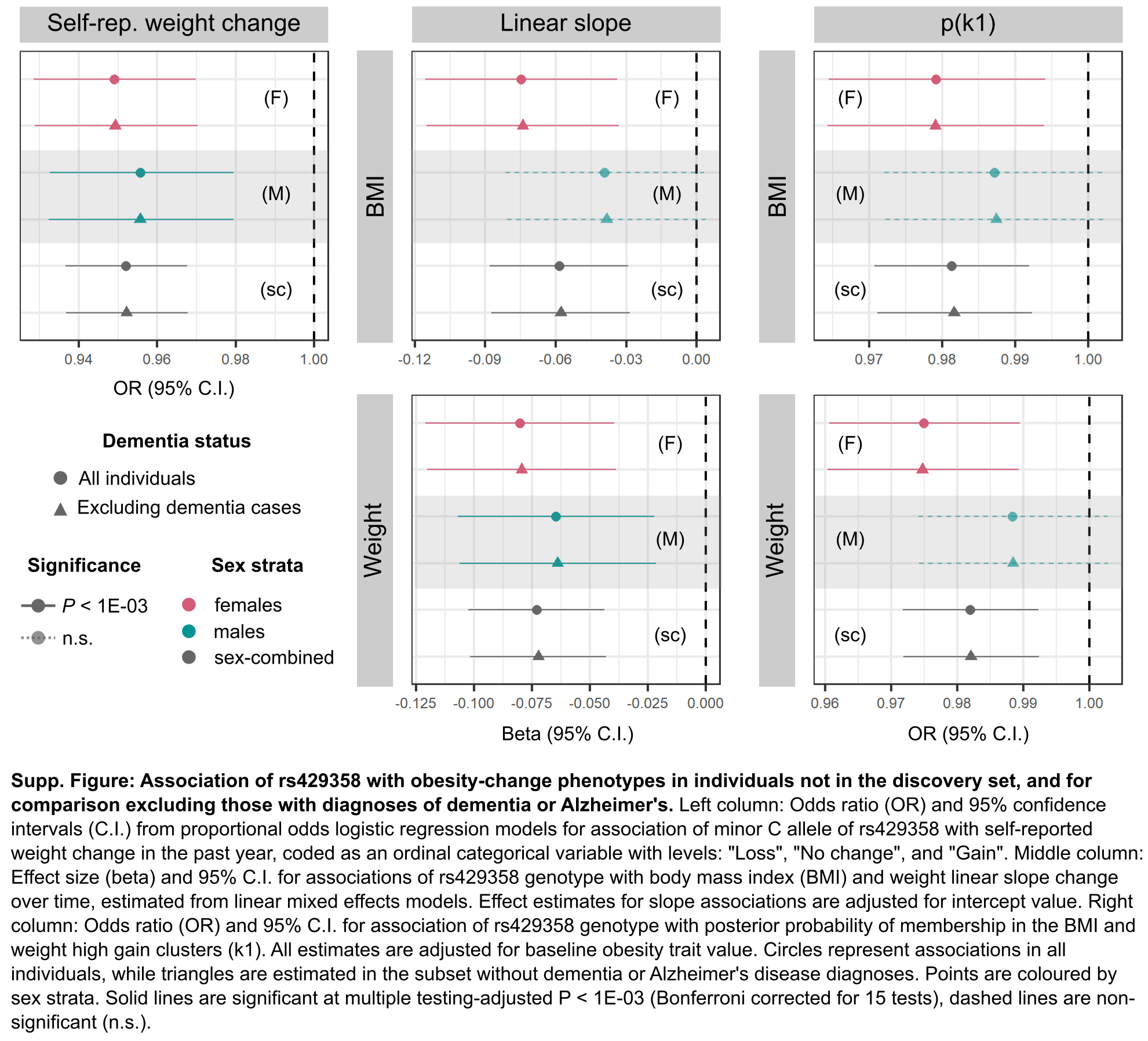

### Supp-Fig-3-supp-methods-data.png

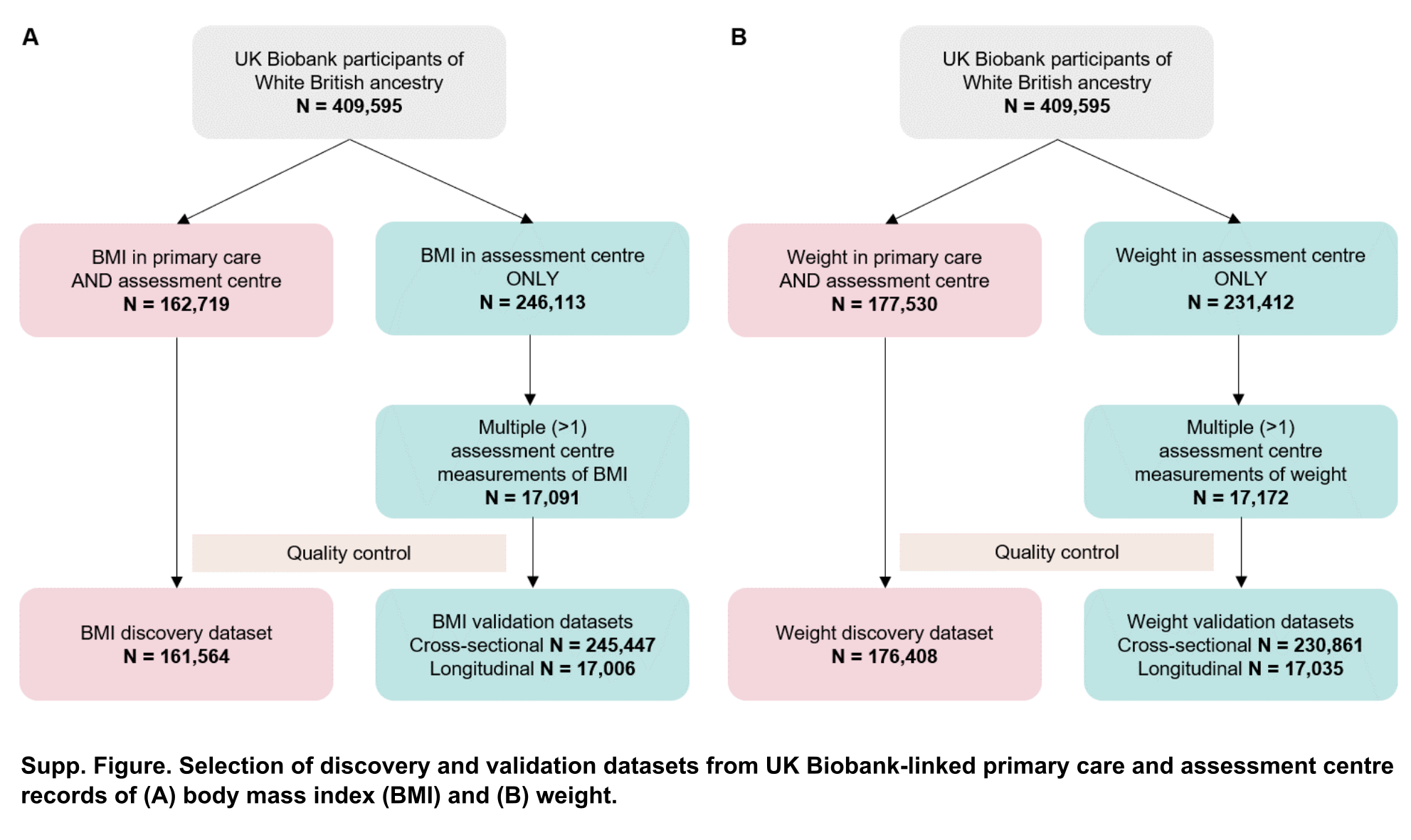

### Supp-Fig-4-supp-methods-ar1-hyperparameters.png

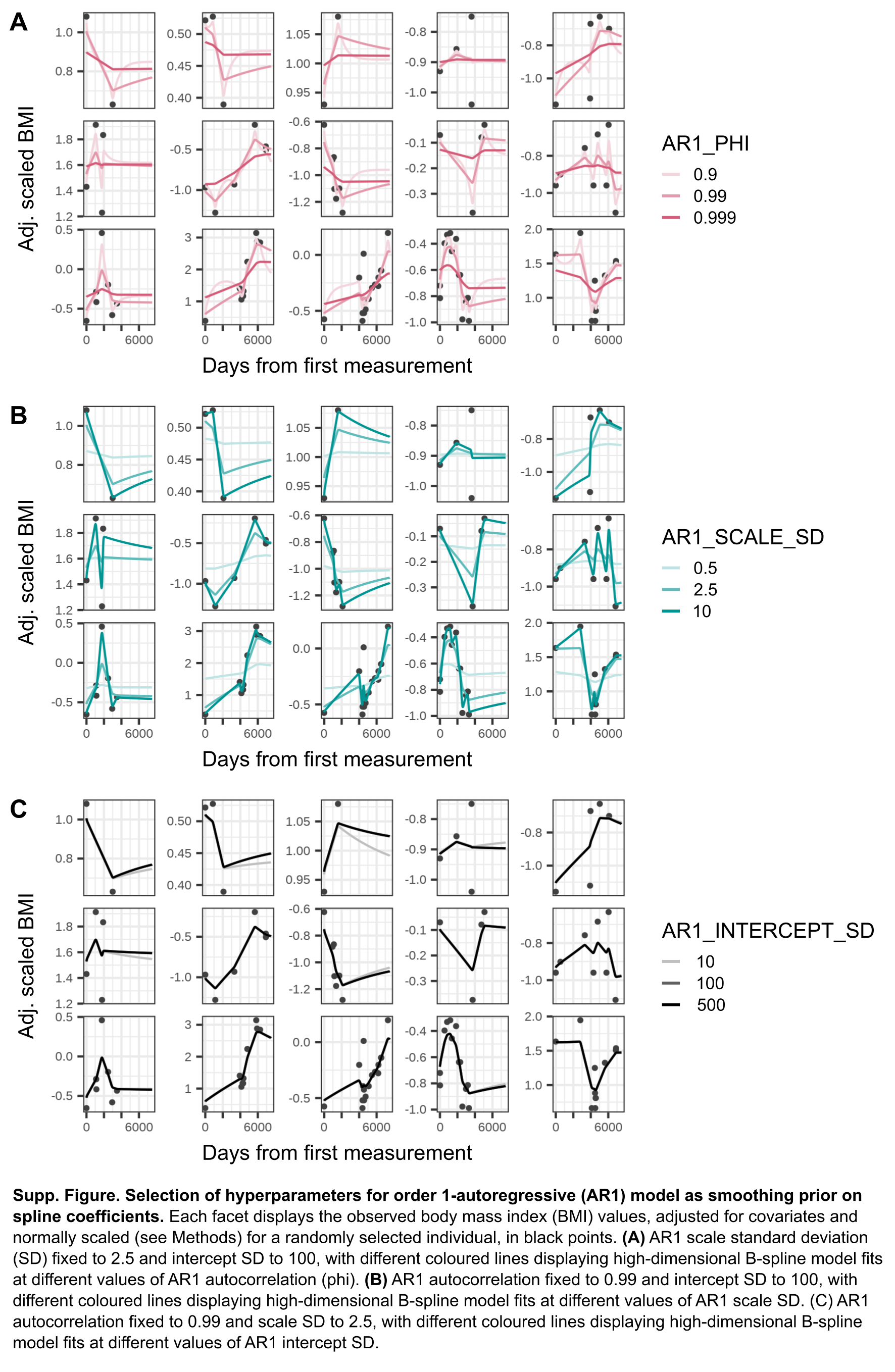

### Supp-Fig-5-supp-methods-clustering-overview.png

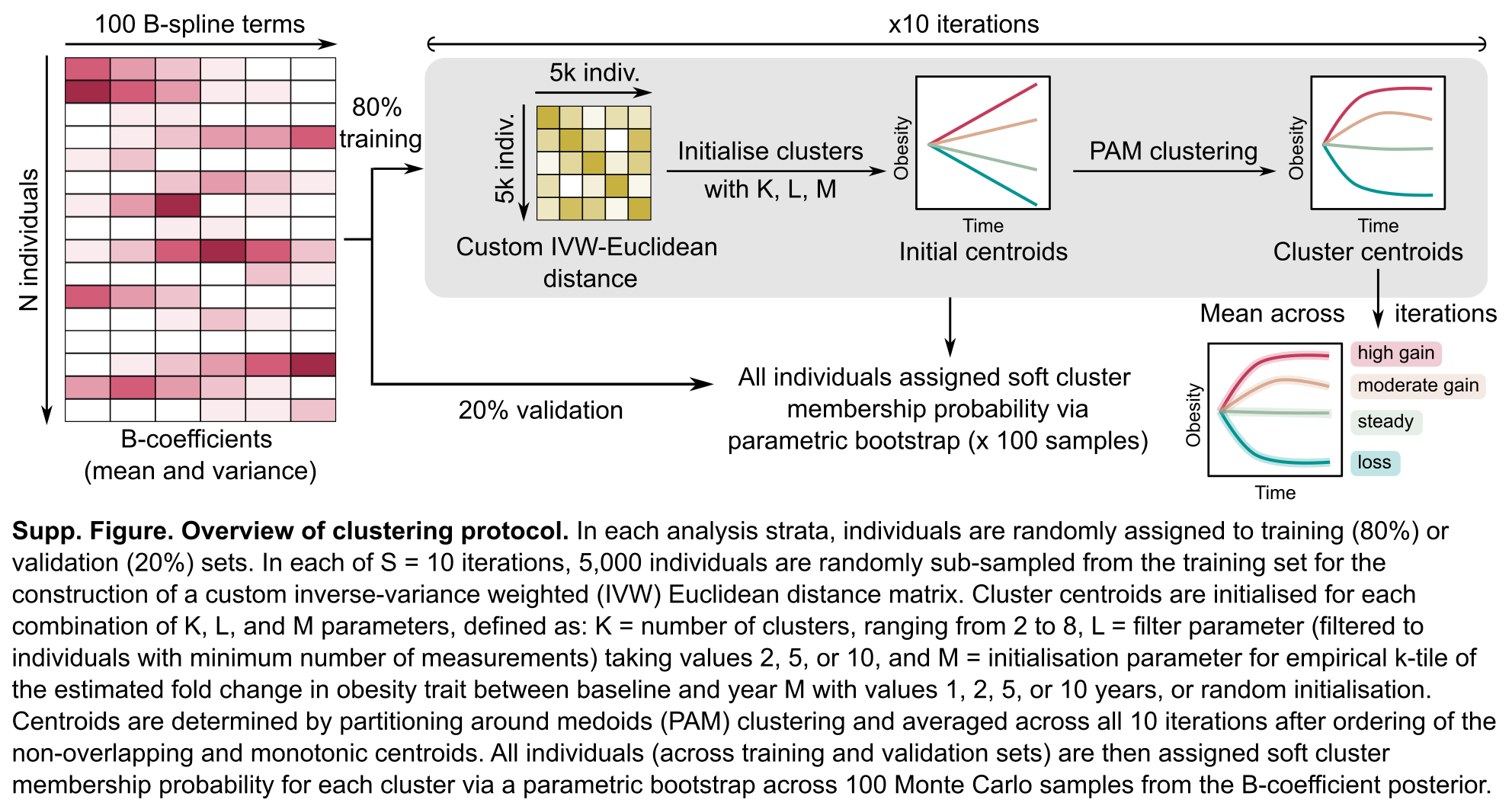

### Supp-Fig-6-supp-methods-cluster-centroids.png

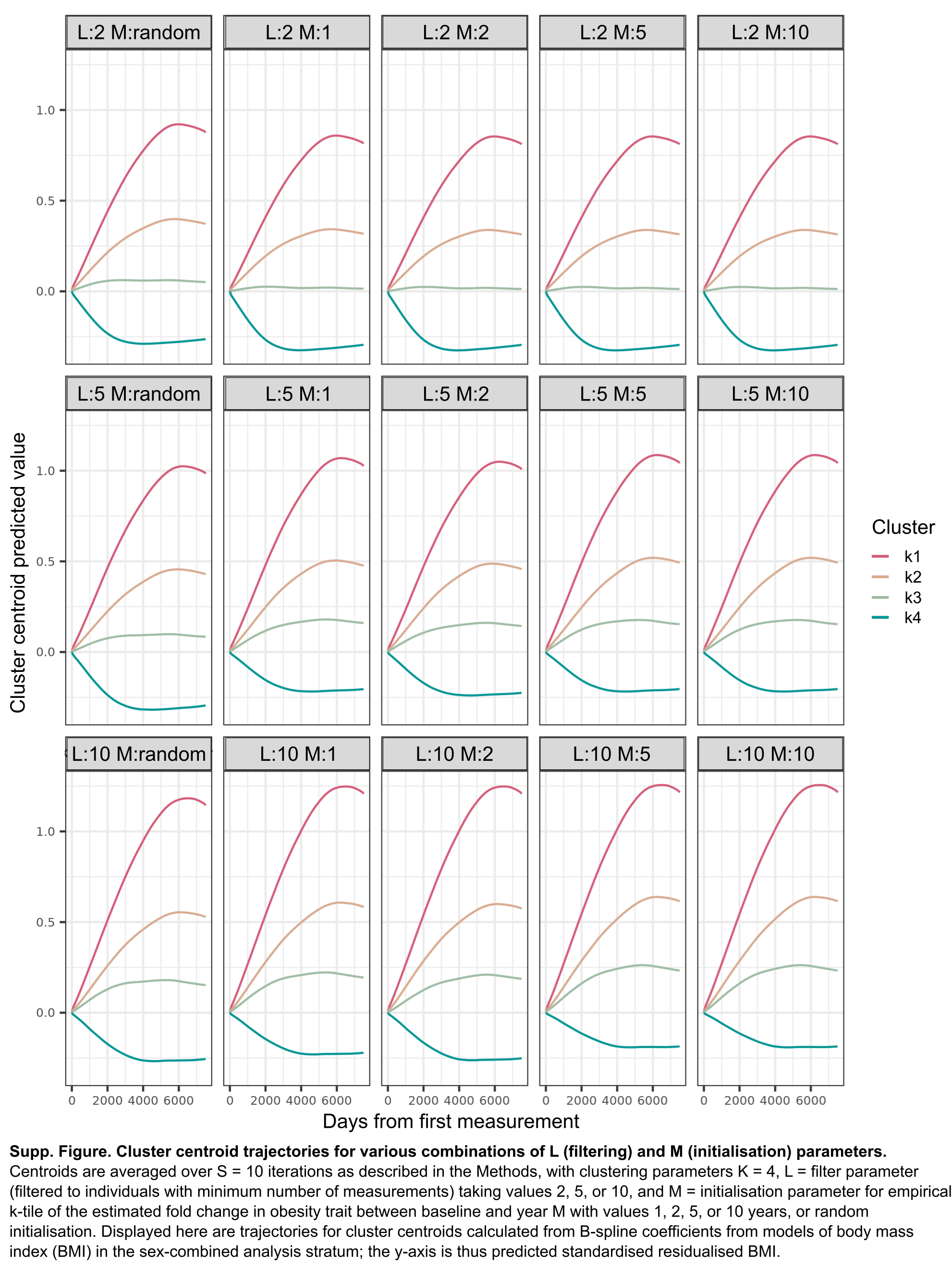

### Supp-Fig-7-supp-methods-klm-silhouettes.png

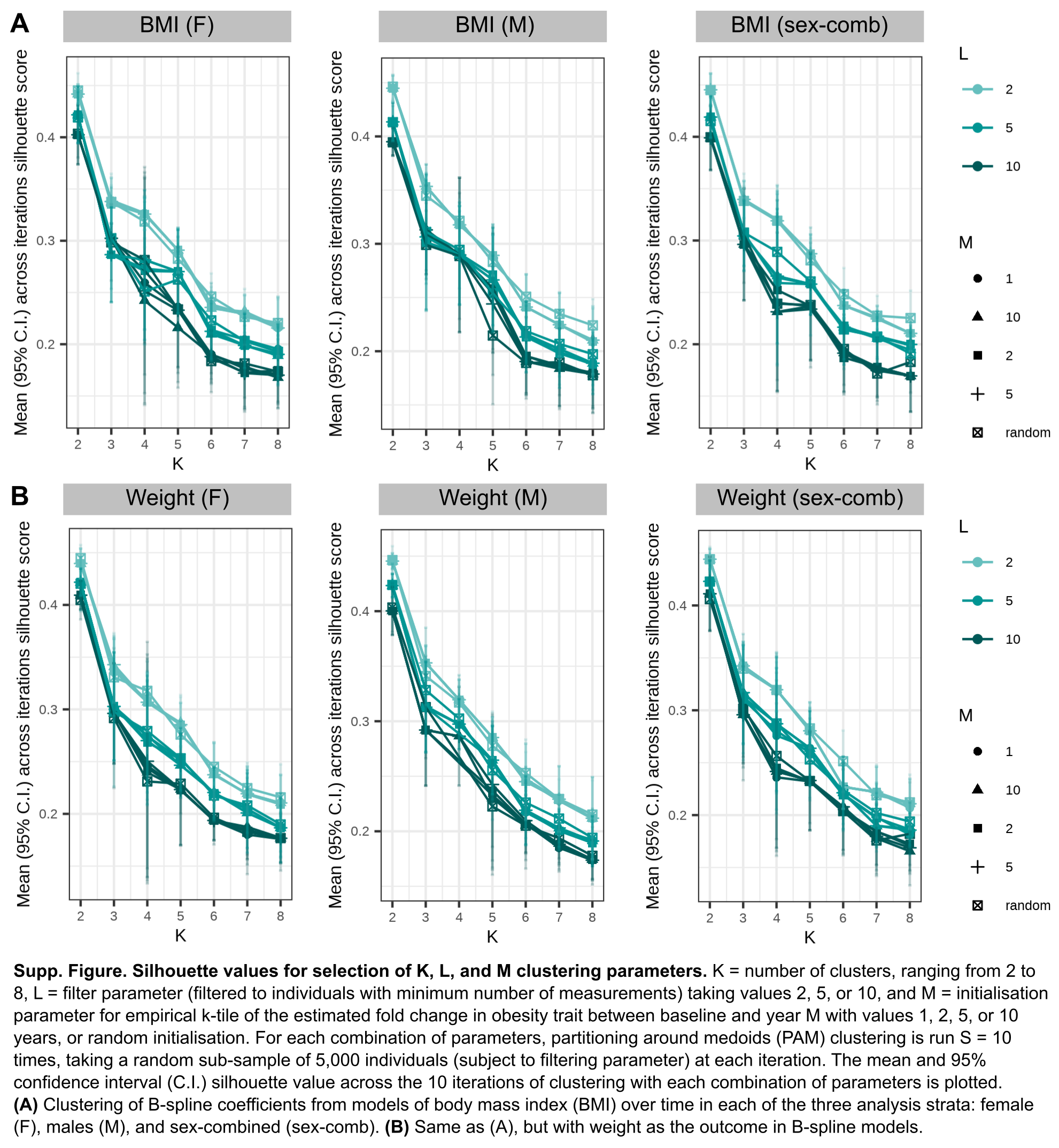

### Supp-Fig-8-supp-methods-clustering-sensitivity-ar1-choices.png

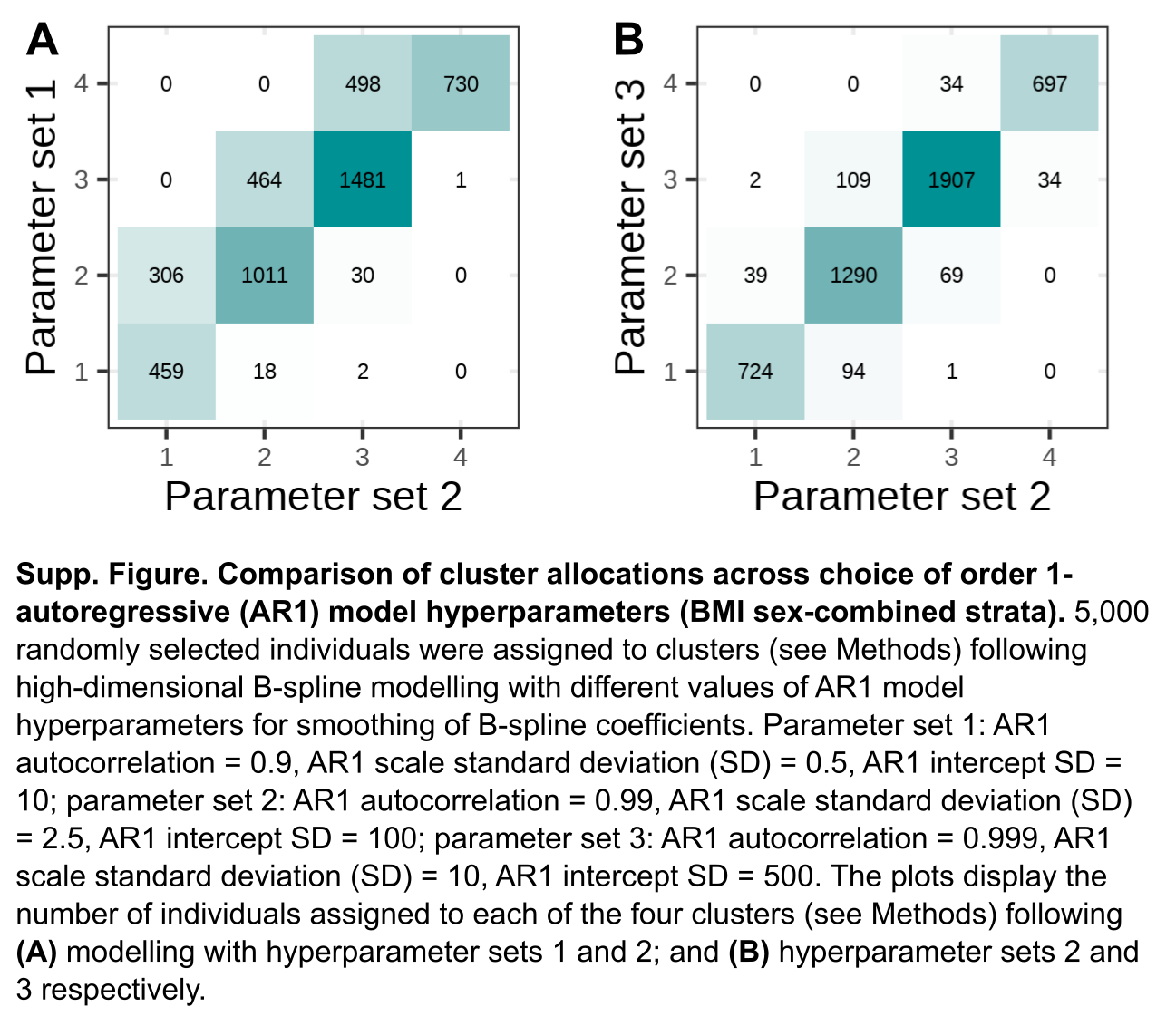

### Supp-Fig-9-supp-methods-clustering-sensitivity-random-train.png

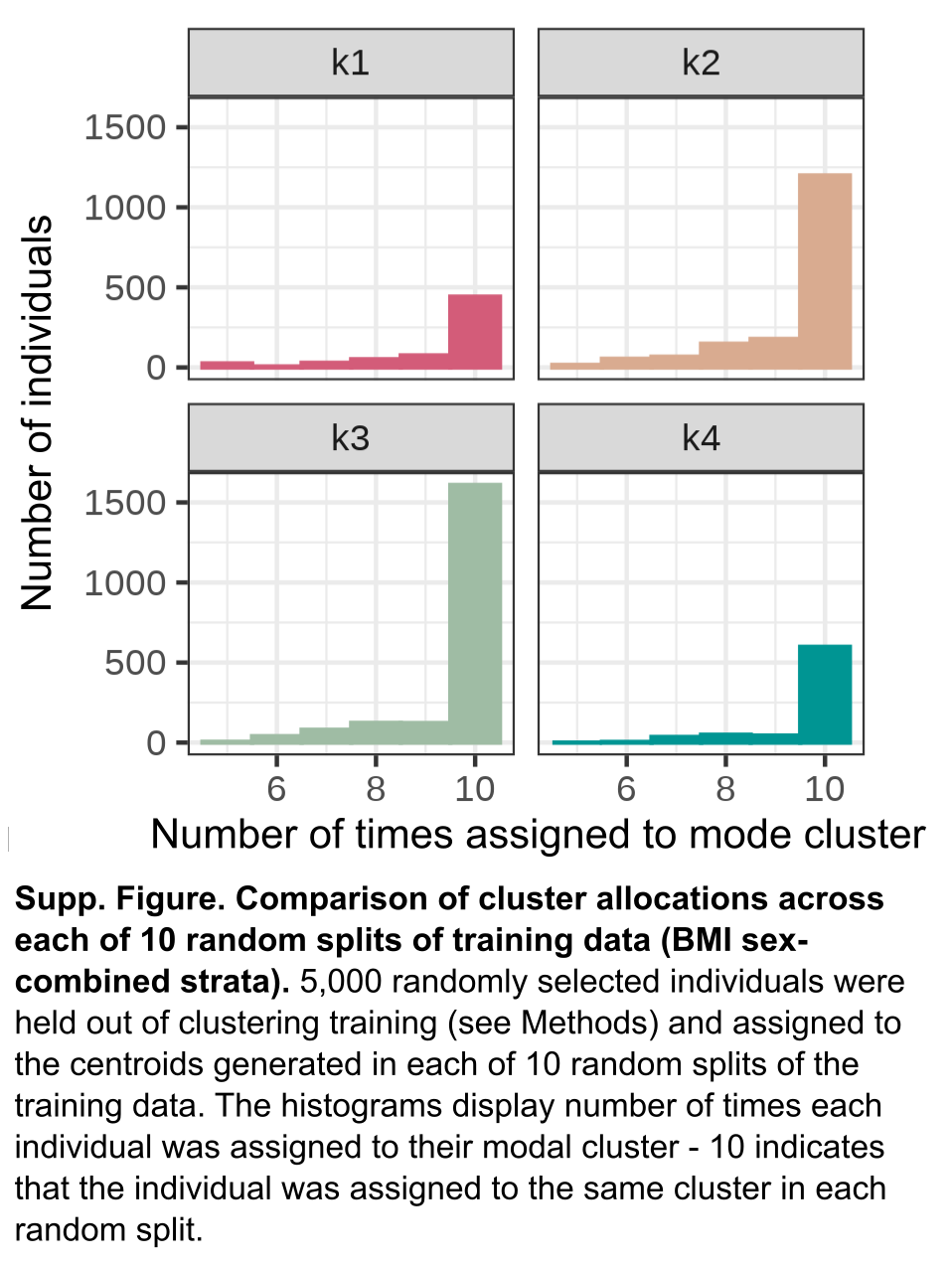
